# A Data-Driven Sliding-window Pairwise Comparative Approach for the Estimation of Transmission Fitness of SARS-CoV-2 Variants and Construction of the Evolution Fitness Landscape

**DOI:** 10.1101/2024.07.23.24310886

**Authors:** Md Jubair Pantho, Landen Alexander Bauder, Sophia Huang, Hong Qin

## Abstract

Estimating the transmission fitness of SARS-CoV-2 variants and understanding their evolutionary landscape is important for epidemiological forecasting. Existing methods are often constrained by their parametric natures and do not satisfactorily align with the observations during COVID-19. Here, we introduce a sliding-window data-driven pairwise comparison method, Differential Population Growth Rate (DPGR), that uses viral strains as internal controls to mitigate sampling biases. DPGR is applicable in time windows in which the logarithmic ratio of two variant subpopulations is approximately linear. We apply DPGR to genomic surveillance data and focus on Variants of Concern (VOCs) in multiple countries and regions. We found the log-linear assumption of DPGR can be found in appropriate time windows in many regions. We show that DPGR can provide estimates of transmission fitness and quantify the transmission advantage of key variants such as Omicron by location. We show that DPGR estimates agree with other methods for estimating pathogenic transmission. Furthermore, DPGR allowed us to construct viral fitness landscapes that capture the evolutionary trends of SARS-CoV-2, reflecting the relative changes of transmission traits for key genotypic changes represented by major variants. The straightforward log-linear regression approach of DPGR may also facilitate its easy adoption. This study shows that DPGR is a promising new tool in our repertoire for addressing future pandemics.

## Introduction

The COVID-19 pandemic caused by the Severe Acute Respiratory Syndrome Coronavirus 2 (SARS-CoV-2) has underscored the importance of estimating transmission fitness for variants to predict the viral evolutionary dynamics. Since December 2019, SARS-CoV-2 has undergone many mutations and likely rounds of recombination, leading to the emergence of multiple variants with varying levels of transmissibility and virulence ^1–3^. Some variants became more dominant during the transmission and were labeled “Variant of Concern” by Greek letters, such as Alpha, Delta, and Omicron ^4–6^.

The basic reproductive number R_0_ is often used to gauge viral transmission, representing the average number of new infections caused by an infected individual in a susceptible population ^7^. R_0_ is often derived from the SIR (Susceptible-Infectious-Removed) model and its many derivations. In practice, the effective reproductive number, R_t_, is often used for a real-time indicator that estimates the average number of secondary infections caused by an infectious individual in a population over time ^8,9^. Both R_0_ and R_t_ are estimated from incidence data, which lack information on particular variants. Consequently, it is often challenging to use R_0_ and R_t_ to differentiate the transmission fitness of emerging variants.

Phylogenetical analysis, as a classic evolutionary approach, has been used to understand the evolutionary fitness changes of SARS-CoV-2 due to its mutations^3,10,11^. Phylogenetic approach has limited usage for SARS-CoV-2 due to the low genetic variability among SARS-CoV-2 sequences, root placement uncertainty, recurrent mutations, recombination among subvariants, and geographic and temporal biases ^10–12^. The limitation of phylogenetic analysis has been highlighted by the unexpected evolutionary pattern of the Omicron variant ^13^.

Multinomial or hierarchical logistical regressions implemented in softmax is a recent approach to estimating the fitness of variants of SARS-CoV-2^14–17^. Multinomial logistical regressions can be used in conjunction with a hierarchical Bayesian framework. The estimated variant fitness can be used for more accurate forecasts up to 30 days and to identify mutations associated with fitness gains.

Alternative approaches for variant fitness estimation can be complementary and address the limitations of existing methods. To this end, we present a sliding-window data-driven pairwise comparative approach to estimate relative fitness for two viral strains, termed Differential Population Growth Fitness (DPGR), based on straightforward log-linear regression. DPGR is an additive distance, and its comparative nature made it more tolerant to some sampling biases. We apply DPGR to estimate the transmission fitness of variants of SARS-CoV-2, construct a fitness landscape for the evolving variants, and shed light on the recent evolution of SARS-CoV-2.

### Differential Population Growth (DPGR) for Pairwise Viral Transmission Fitness Comparison

Motivated by using an internal control to mitigate sampling biases often associated with SAS-CoV-2, we designed a pairwise comparative approach on sliding time windows, the Differential Population Growth Rate (DPGR), for estimating the pairwise transmission fitness advantages of the variants of SARS-CoV-2. DPGR is calculated by log-transforming the ratio of the growth rate of two exponentially growing populations in the applicable time windows and taking the growth latencies into account, as shown by Equation 1.

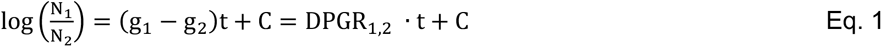

where N_1_ and N_2_ are two exponentially growing viral sub-populations, g_1_ and g_2_ are the growth coefficients, and the constant C incorporates the lag time of the growth of the two sub-populations, and t represents time which is measured in days in this study. The log-transformed ratio of the growth is a linear model where the positive slope indicates population N_1_ grows faster than population N_2,_ and the negative slope indicates the opposite. DPGR_1,2_ = (g_1_ - g_2_) is defined for variants 1 and 2. (Detailed induction of DPGR is in the Supporting Document.) In practice, sliding time windows were applied, and the appropriate periods for the log-linear approximation were selected based on linear fitting.

During COVID-19, some variants dominate in different periods and are challenging to be co-sampled at the same location in the same time window, especially given the limited genomic surveillance capacity. Based on the property of logarithms log(a/b) = log(a/c) + log(c/b), we can find another variant c which can estimate DPGR_a-b_ through DPGR_a,b._ = DPGR_a,c_ + DPGR_c,b_ as long as variant c can be co-sampled with both variant a and b at the same location. (See detailed formulation in the supporting documents.) For instance, we estimated DPGR_Alpha-Delta_ through DPGR_Alpha,Delta_ = DPGR _Alpha,Beta_ + DPGR _Beta,Delta_, even though we could out find sufficient weekly co-observations of Alpha and Delta variants in the same location in the genomic sampling data from GISAID^18^.

## Results

### Overview of the Computational Flow Work

As shown in Figure 1, we retrieved the genomic variant surveillance data for SARS-CoV-2 from GISAID up to June 16, 2022. The Variant of Concerns (VOCs) in the retrieved data set include Alpha, Beta, Gamma, Delta, and Omicron. The GISAID surveillance data include collection dates and locations. We calculated the weekly average occurrence of each VoC at selected locations and scanned for time windows in which DPGR is applicable. The DPGR-applicable time windows were typically selected for at least 4 weeks and with a linear fit of R^2^ value greater than 0.9. We applied DPGR to various countries and continents. For comparisons, we estimated the transmission fitness for VoCs designated by WHO and the Pango lineage^19^ provided by GISAID. Based on the pairwise fitness estimation, we constructed fitness stairs and landscapes to capture the evolutionary big picture of SARS-CoV-2.

**Figure 1.**
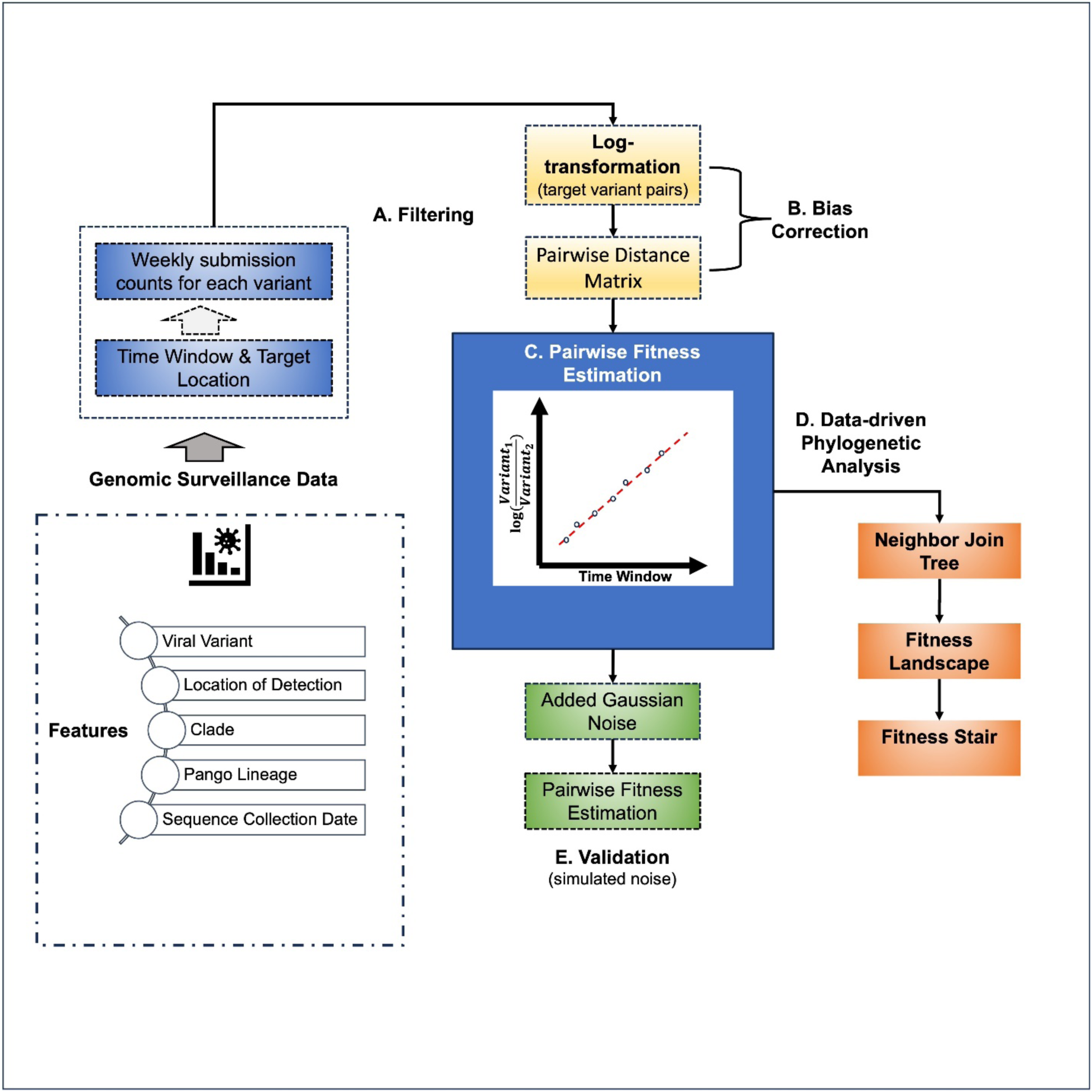
Overall Computational Workflow of the DPGR Model for Transmission Fitness Estimation.

### Country Level Viral Transmission Revealed by DPGR with Omicron and Delta as an Example

We first examined the utility of DPGR to estimate variant transmission fitness at the country level using the pair of Omicron and Delta variants as an example. We performed sliding time window analysis to select time windows in which the DPGR assumption held well. As an example, estimations of DPGR between Omicron and Delta in a few countries are presented in Figure 2. A set of target countries, including the United States, Canada, Brazil, South Korea, Ireland, Denmark, Netherlands, Italy, Turkey, Belgium, Poland, Israel, Japan, Switzerland, Spain, France, Mexico, and Germany, are selected for showcasing the application of DPGR for estimating the transmission fitness (See Figure 2 and Figure S1). In the United States, within the time window from March 2022 to May 2022, the logarithmic values of Omicron versus Delta population fit well with a linear model (Figure 2 A) with an R^2^ value of 0.99 (See Table S1 for the estimated slope and R^2^ values for the aforementioned target countries). Applying DPGR in the selected countries yields a range of estimation of Omicron versus Delta, termed DPGR_Omincron,Delta_ (Figure 2G), ranging from 0.008 to 0.1 with the average at 0.06. Among the analyzed countries, the highest DPGR_Omincron,Delta_ is observed in Turkey (0.1) and the lowest in the United States (0.08). It can be observed that (see Figure 2 G) that some European bordering countries (Denmark, Germany, Belgium, etc.) exhibited similar DPGR_Omincron,Delta_ - indicating that these bordering countries share similar trends of SARS-CoV-2 epidemic transmission based on DPGR estimates.

**Figure 2:**
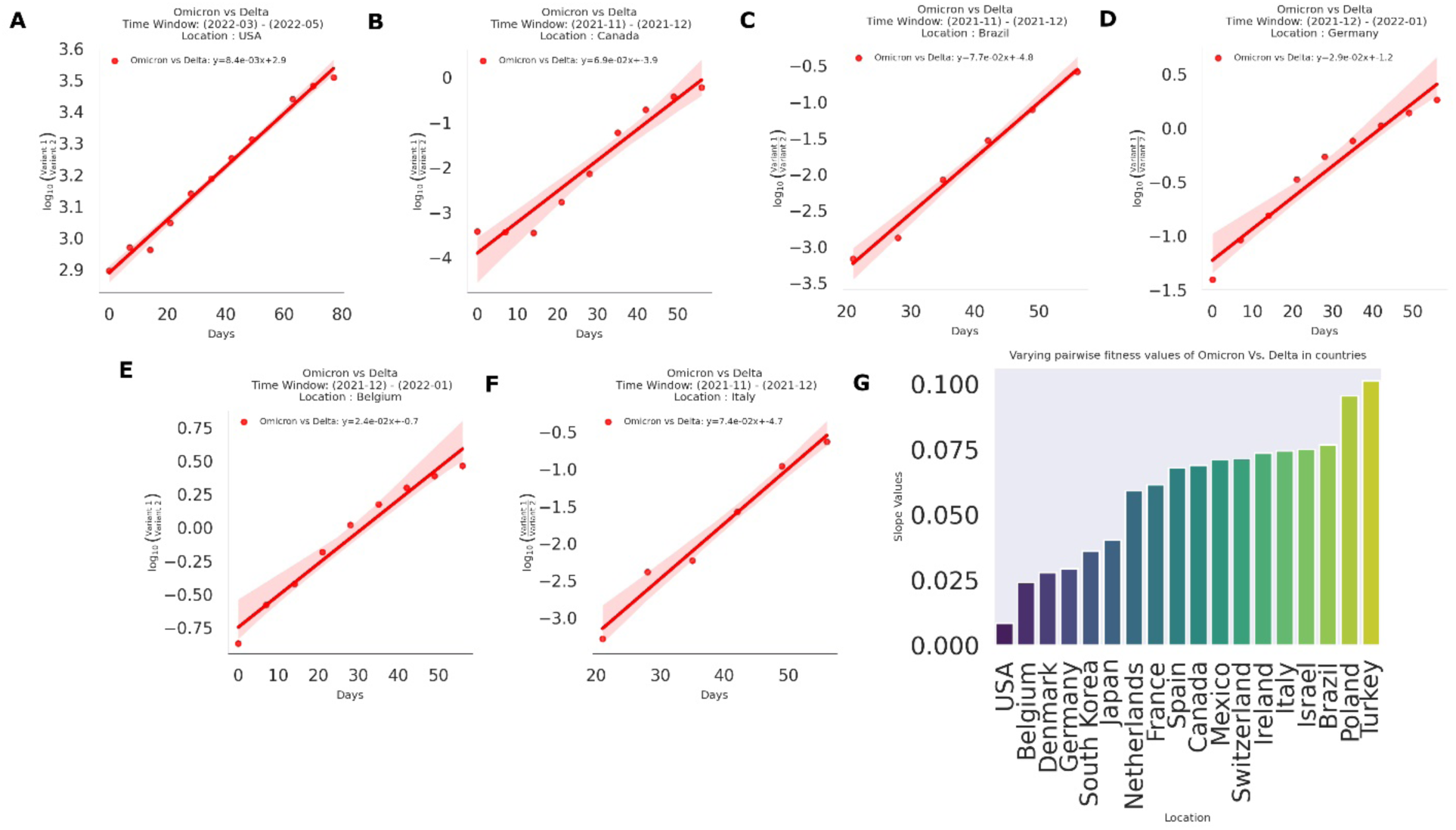
Pairwise Transmission fitness estimation of Omicron compared to Delta in several countries. The figures (A-F) Illustrates the sharp increase of pairwise transmission fitness of Omicron Compared to Delta in the target countries. The y-axis in the plots refers to the log-transformed ratio of growth, and the x-axis refers to the time window in days. G. The bar plot visualizes the estimated transmission fitness values in all the analyzed regions.

As a comparison, we also fit DPGR to the Pango lineage labels GRA and GK, which correspond to the WHO labels of Omicron and Delta. The plots in (see Figure S3) give a visual representation of the models’ estimation for GISAID labels. The estimates, designated DPGR_GRA,GK_ are nearly identical to DPGR_Omicron,Delta_ based on WHO labels. These results demonstrate the generalizability of DPGR. The pairwise transmission fitness values between the variant pairs of both WHO and GISAID labels are illustrated in the heatmap (see Figure S2 and Figure S4). By applying DPGR for the target countries, the observed DPGR_GRA,GK_ ranges from 0.0094 (USA) to 0.1021 (Turkey) with R^2^ values of 0.99 and 0.97 respectively, indicating similar estimates as DPGR_Omicron,Delta_ (see details in Table S6).

Overall, the country-level analysis shows that DPGR is useful for examining the country-level variations in viral transmissions, which might reflect differences in regional responses, vaccine efficacies, population demographics, immune responses, environmental factors, and social and cultural factors.

### Continent Level Viral Transmission Revealed by DPGR with Omicron versus Delta variant as an example

To further examine the utility of DPGR, we applied it at the continental level using the Omicron versus Delta variants as an example. We preprocessed the data to weekly frequency for each variant in each continent. Figure 3 (A-G) illustrates a positive rise in transmission fitness advantage of Omicron in several continents in different time windows, which indicates the geographic mobility of the circulating Variant of Concern. The DPGR estimates range from 0.006 to 0.06, with an average value of 0.03. The lowest relative transmission fitness is observed in Oceania (0.03) and the highest (0.06) in Asia and South America, with Africa, North America, and Europe in between. The R^2^ values ranges from 0.92 to 0.99, with the highest R^2^ values observed in North America(0.99), South America(0.99), Oceania(0.99) and Asia(0.99), and the lowest in Africa(0.92) (see Table S1).

**Figure 3.**
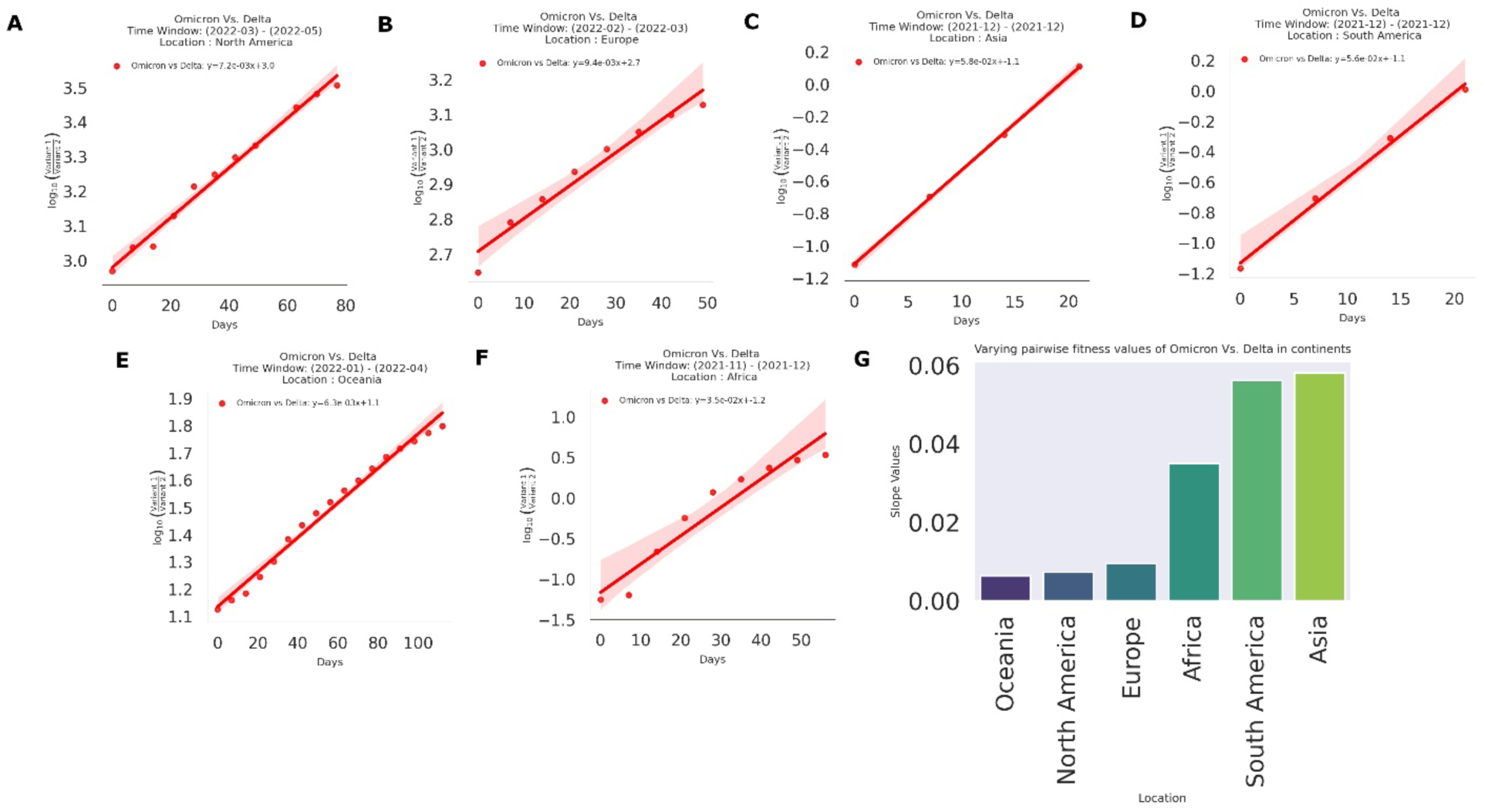
Pairwise Transmission fitness estimation of Omicron compared to Delta in several continents. The scatterplots (A-G) visualize the sharp increase of transmission fitness of Omicron compared to Delta in several continents. G. Bar plots to visualize the fitness values in each continent

We estimate DPGR for the corresponding GISAID labels GRA and GK (see Figure S5) which show nearly identical trends. The pairwise transmission values between the variant pairs (GRA, GK, GRY, and GH) are illustrated as heat maps in Figure S6. The GRA variant demonstrates a relative fitness advantage, particularly in Asia and South America, with the smallest increase in Oceania. The range of the transmission fitness values between the comparison, DPGR_GRA,GK_, in several continents is 0.0064 (Oceania) to 0.0577(Asia) with R^2^ values 0.99(0ceania) and 0.99(Asia) respectively. The estimate between DPGR_GRA,GK_ indicates analogous estimated values as DPGR_Omicron, Delta_ estimates for the continents (Table S6).

Overall, the results here show applying DPGR at different geographic scales can reveal the regional heterogeneity of viral transmission and aid our understanding of the spread and impact of different SARS-CoV-2 variants.

### Transmission Variations of Omicron Sublineages Revealed by DPGR

To further examine the utility of DPGR, we applied DPGR in evaluating the sublineages of the Omicron variant^20,21^. From the Pango Lineage of the GISAID dataset, we selected the sublineage BA.1 ∼ BA.5 associated with the Omicron variant, estimated the weekly frequency for each sublineage at a location of interest, and selected the time windows in which the linear fit of DPGR can be observed. For North America, the R² values for the comparisons between BA.5 and other variants within the time window of April 2022 to May 2022 are as follows: BA.5 vs. BA.1 (0.982), BA.5 vs. BA.2 (0.985), BA.5 vs. BA.3 (0.993), and BA.5 vs. BA.4 (0.931). See Table S2 for details.

For the sake of illustration, we chose the dominant sublineage BA.5 to compare with the other circulating sublineages (BA.1, BA.2, BA.3, and BA.4). Figure 4 (A-E) shows the pairwise DPGR estimation plots of BA.5 with other sublineages in five continents in time windows ranging from 5 ∼ 11 weeks in 2022. The linear fit of DPGR for the Omicron sublineages was often more pronounced than those in comparisons of major variants, such as Omicron and Delta (Figures 2 and 3). The DPGR analysis demonstrates that the Omicron sublineage BA.5 consistently dominated over other sublineages though at varying levels of relative fitness compared to other sublineages (BA.1, BA.2, BA.3, BA.4) across different continents, as summarized in Figure 4F. For example, in Asia, BA.5 was found to have the fastest differential growth advantages over others, whereas its growth advantage was relatively moderate in Africa. In Europe and Asia, the DPGR plots of BA.5 over BA.4 were nearly flat, indicating codominance during the period of the analysis.

**Figure 4:**
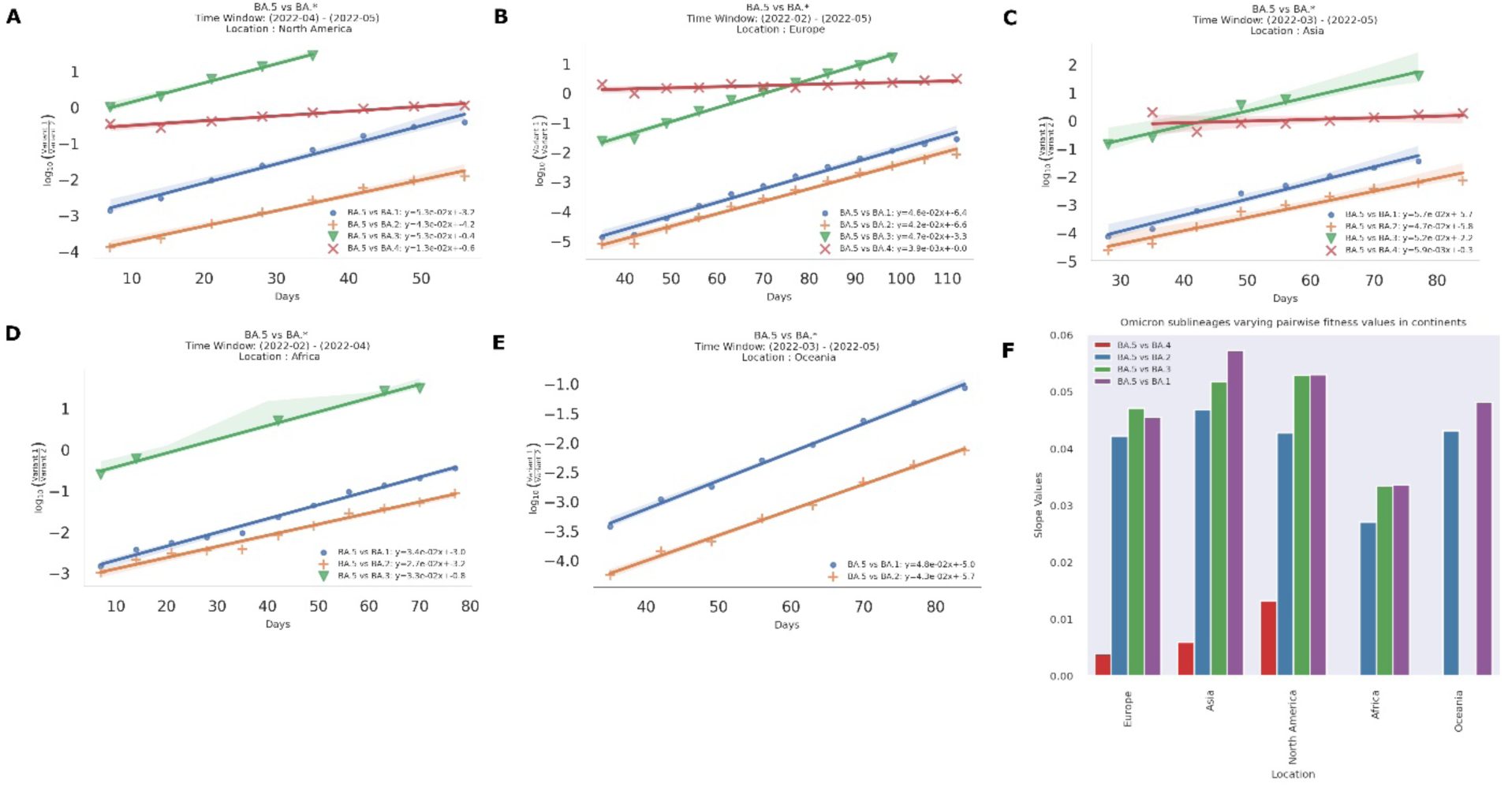
Pairwise Transmission fitness estimation of Omicron sub-lineages in target continents. The subplots(A-E) Illustrates the estimated pairwise transmission fitness of Omicron Sub-lineages (BA.5 with others sublineage). F. estimated fitness values in several continents.

### The Evolution Fitness Landscape of SARS-CoV-2 Variants and Sub-variants

The pairwise approach of DPGR offers an opportunity to capture the relative fitness landscape during the evolution of SARS-CoV-2 variants. At the time of our study, we have data to address the Alpha, Beta, Delta, and Omicron VoCs in the United States. All possible pairwise comparative analyses among these four VoCs lead to a 4x4 matrix (see heatmap of Figure 5A). Examination by sliding windows shows DPGR model is applicable to Alpha and Beta from March 2021 to April 2021, to Beta and Delta from August to September 2021, and to Delta and Omicron from March to May 2022 with corresponding R^2^ values of 0.98, 0.97, and 0.98 respectively. For example, DPGR_omicron,beta_ = 0.015, and DPGR_beta,omicron_ = -0.015, which suggests that Omicron out-grow Beta by 1.5% daily (because e^0.015^ »1.015). Some pairs of VoCs, such as Alpha and Delta, though do not have sufficient data points due to non-overlapping periods, can be inferred based on the property of logarithms log(a/c) = log(a/b) + log(b/c) (See details in the supporting documents), which enables us to compare transmission fitness for variants that dominate in different periods during the pandemic. For instance, we estimate DPGR_Alpha,Delta_ = DPGR _Alpha,Beta_ + DPGR _Beta,Delta_, even though we could out find sufficient weekly co-observations of Alpha and Delta variants in the same location in the genomic sampling data from GISAID.

**Figure 5:**
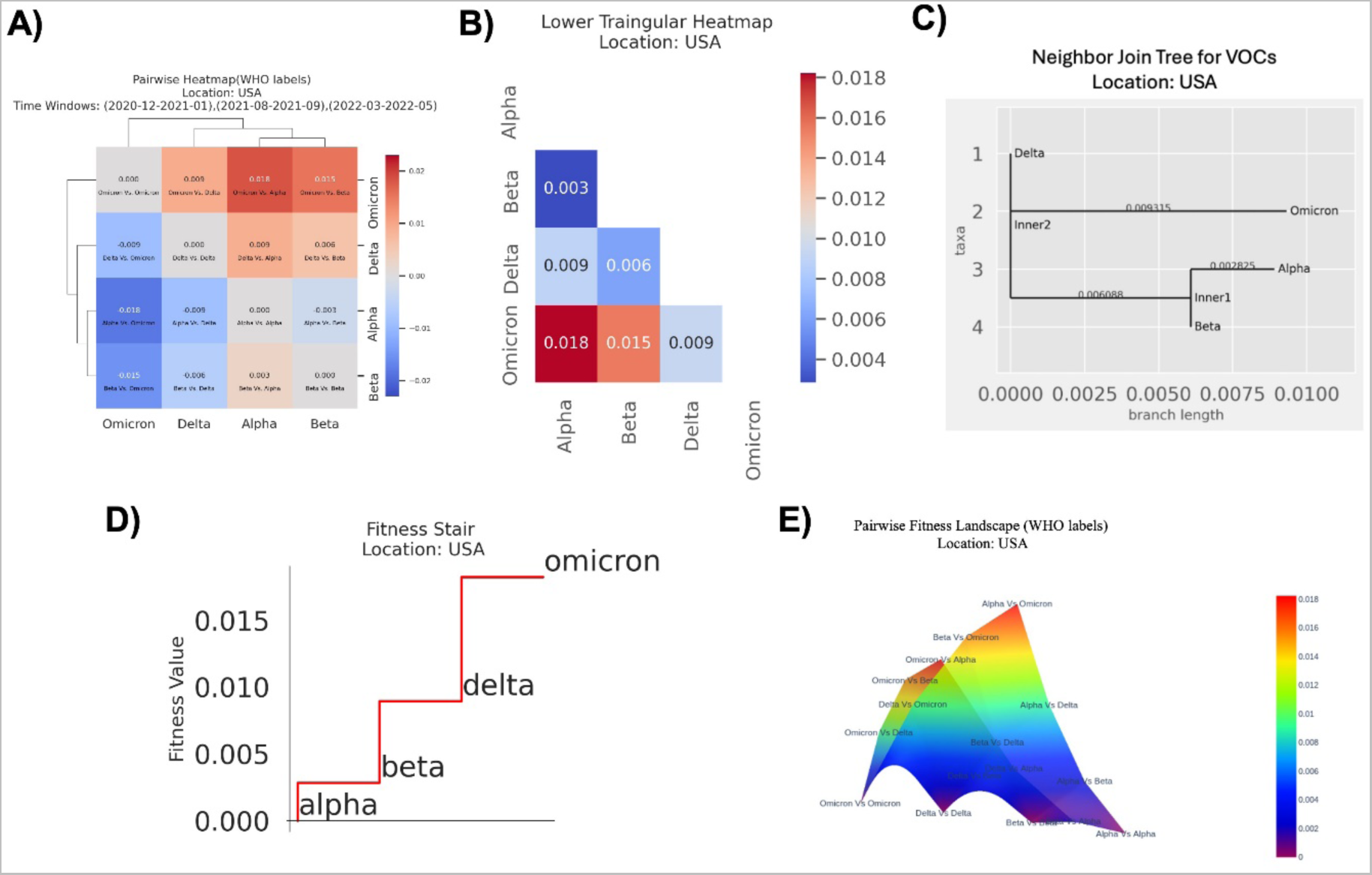
Progressive fitness evolution of Variants of Concerns (VOCs) a) Pairwise heatmap of the transmission fitness for WHO variants in USA. B) Lower triangular heatmap with absolute pairwise distance to create the NJ tree, Fitness Stair, and Fitness Landscape. c) Neighbor Join tree d) Step plot visualizes the progressive transmission fitness gain of the WHO labeled Variant of Concerns (VOCs) in the United States. e) illustrates the pairwise transmission fitness landscape of the SARS-CoV-2 variants in USA for the WHO labels. Variant Pairs having higher pairwise transmission fitness is on the top of the landscape and vice-versa.

It is worthy of emphasizing the DPGR is a relative measurement, and it is often more convenient to present the fitness on an ‘absolute’ scale. To do this, we first convert the DPGR matrix into a distance matrix (see Figure 5B) by only focusing on the positive values. From the distance matrix, we apply the neighbor-join tree^22^ to capture the representative tree structure of fitness for the variants (see Figure 5C). From the neighbor-join tree, we infer the transmission fitness on a scale using the initial VoC Alpha as a reference point.

We inferred a fitness staircase (see Figure 5D) depicting the transmission fitness gain of the VoCs — Alpha, Beta, Delta, and Omicron—in the United States. The progressive transmission fitness values for Beta, Delta, and Omicron are shown relative to the initial variant, Alpha. The substantial increase in transmission fitness from Delta to Omicron is clearly illustrated by the fitness staircase.

We constructed a neighbor-join tree for Omicron sub-lineages (BA.1*, BA.2*, BA.3*, BA.4*, BA.5*) to gain a closer understanding of the progressive fitness evolution (Figure S12). We found that BA.5 has the fastest growth advantage with BA.4 as a close competitor. BA.2 and BA.4 have nearly similar fitness; however, BA.5 has rapidly increased transmission fitness than the other two sub-lineages. Collectively, BA.5 and BA.4 caused a rapid surge of cases in the United States likely due to their escape of neutralizing antibody^20,21,23^.

We constructed a fitness landscape for these circulating variants in the U.S.A. (Figure 5E) which illustrates the relative change of transmission fitness among the variants during their evolution. The fitness landscape in evolutionary biology is used to visualize the relationship between genotypes and their evolutionary fitness^24,25^, and is of importance to understanding viral evolution^26–31^. In Figure 5E, we use the variants to represent the change of genotypes of SARS-CoV-2. The flying bird-shaped landscape (See Figure 5E) visualizes the pairwise fitness landscape of the WHO-labeled Variants of Concern (VOCs). To generate the fitness landscape, we used the absolute pairwise DPGR. The pairs with the lowest fitness values are presented at the bottom of the landscape, and those with higher fitness climb progressively on the top of the hill. At the bottom of the hill Omicron Vs. Omicron has zero fitness (from the estimated pairwise distance matrix), and as moved uphill, Omicron exhibits significant fitness gain against Delta, Beta, and Alpha. At the top of the hill, Omicron Vs. Alpha has the highest fitness gains as shown by the highest value of DPGR between the Omicron and Alpha. The Delta variant can be seen in the middle of the hill, reflecting its relative growth advantage between Omicron and other variants.

Overall, we show that the pairwise approach of DPGR can allow us to infer the fitness distance matrix fitness landscape, and fitness staircase through the neighbor-join tree method.

### Comparison of DPGR with other measurement

We compared the relative transmission fitness of SARS-CoV-2 variants using the Differential Population Growth Rate (DPGR) model and the PyR_0_ model developed by another study^14^. We performed a linear correlation analysis (see Figure S 13) between the estimated DPGR and those by PyR_0_. We used the fold increase in relative fitness (R/RA) for the Alpha, Beta, Delta, and Omicron variants, relative to the Wuhan A strain, as provided in the “strains.tsv” file from the supporting materials of the previous study. We log-transformed (log2) the fold increase to make the values comparable with the DPGR model. We observed a strong linear correlation (R^2^ = 1) between the estimates of PyR_0_ and DPGR. Interestingly, the requirement of a log transformation shows that the estimates of DPGR and PyR_0_ are on different scales. These analyses show that on one hand, DPGR gave a comparable estimate to the relative fitness of SARS-CoV-2 variants, and on another hand, DPGR offers a different perspective to the previous method.

## Discussions

In this study, we present a data-driven sliding-window pairwise approach, DPGR, to capture the relative competitiveness of SARS-CoV-2 subpopulations and their changes in fitness during the pandemic. The pairwise nature of DPGR allows one subpopulation to serve as an internal control. The simplicity of log-linear regression can facilitate the wide implementation and adoption of DPGR. The additive property of DPGR makes it suitable for distance matrix-based analysis. Limitations of DPGR include the exponential growth assumption and the need for correctly labeled variants. To avoid the negative value of DPGR, the dominant variant should be chosen to be the numerator part of DPGR.

In contrast to the typical parametric fitting approach of the existing methods, the sliding time windows approach focuses on the most applicable period for DPGR. Our studies show that these applicable time windows are readily available during the COVID-19. The pairwise ratio approach can help reduce sampling errors that are non-discriminatory to subvariants. Surveillance data on SARS-CoV-2 genomics are considerably under-reported. It is reasonable to argue that most genomic surveillance methods are non-discriminatory with respect to viral genomic sequences. Hence, the ratio between the two observed measurements would cancel out the non-discriminatory sampling errors. We acknowledge that this pairwise approach assumes the subpopulation clustering is correct and equal sampling among the subpopulations. It is important to discuss the similarities and differences between DPGR and the classical logistical growth model^3^. DPGR is different from the logistic growth model log(p_1_/(1-p_1_)), in the sense that the denominator was replaced by the fraction of another subpopulation p_2_. Hence, DPGR can accommodate the heterogeneity of relative fitness in subpopulations as observed in the sub-populations of Omicron.

Notably, DPGR_Omicron_Delta_ in the USA is the smallest compared to those in European countries. Regional differences in DPGR may be influenced by various factors. For example, age-dependent infection rates of variants^32^ and human population age structures could contribute to these regional differences. Variants’ differential dependence on environmental factors^33^ such as humidity and temperature is another possibility. Additionally, viral mutations^6,34,35^, host genetic makeup^36^, immune responses^23,37^, and drug resistance^38,39^ may play a role. Variations in vaccine efficacies are also a contributing factor.

Finally, the estimated fitness and relative changes in fitness of SARS-CoV-2 variants would be a valuable resource for genome-wide association studies, which can help identify mutations associated with fitness changes in SARS-CoV-2 and interactions with host genetic factors.

## Materials and Methods

### GISAID Meta Information

The dataset used for this research is provided by GISAID^18^, which stands for Global Initiative on Sharing All Influenza Data. We download the metadata tsv file. There are a total of 18 columns in the dataset. We mostly focused on Location, Clade, Pango Lineage, Variant, and Collection date. The variant column records the type of the SARS-CoV-2 variant according to the WHO (World Health Organization) proposed labels. Only Variants of Interest (VOI) or Variant of Concern (VOCs) are labeled by WHO.

Pango Lineage is a dynamic nomenclature system that tracks the transmission and spread of the SARS-CoV-2 variants. Pango Lineage uses phylogenetic diversity for naming the SARS-CoV-2 lineages that contribute most to the current propagation.

#### Variant of Concern (VOC) is defined by WHO

When a variant of newly emerged SARS-CoV-2 changes the pandemic dynamics and causes increased hospitalization and death, the World Health Organization (WHO) labels that specific variant as a Variant of Concern (VOC). WHO also keeps the preventive measures on alert to check the further propagation. There are five variants of Concern (VOCs) labeled by WHO: Alpha, Beta, Gamma, Delta, and Omicron.

### Data Preprocessing

In the data preprocessing stage, we have taken some steps to ensure the data is in the correct format before passing it to the DPGR model. Initially, the raw dataset is loaded from a TSV (Tab-separated value) file using libraries from pandas. Filtering of the rows is performed to ensure that the rows containing the non-null ‘Variant’ values are selected. We also mapped the variant name to a standardized format aligning with the GISAID or Pango nomenclature system to ensure consistency in variant names along the entire dataset. This ensured the uniformity of the naming of all the variants. Depending on the region of interest (country or continent), the ‘Location’ column is mapped to the predefined lists of regions to facilitate country or continent-level analysis. This standardization facilitated more accurate geographical analysis. Subsequently, the dataset was narrowed down to the most pertinent columns: ’Variant’, ’Location’, and ’Collection date’. The ’Collection date’ was then converted to a Python datetime format, and a new column representing the collection week was introduced to enable the weekly aggregation of the data.

The dataset is further filtered to a specified date range (’2020-01-01’ to ’2022-05-31’) and grouped by ’Variant’, ’Location’, and ’Date’ to obtain weekly frequency counts for a certain variant at a target location. Certain variants, which were less relevant for the analysis, were excluded to focus on the primary variants of interest such as Alpha, Beta, Gamma, Delta, and Omicron. The ’Date’ column was refined to extract the start date of each week, ensuring clarity in temporal analysis.

This weekly dataset was further summarized by summing the frequencies of each variant for each week and location, providing a clear and concise dataset for further analysis. This comprehensive preprocessing stage ensured the dataset was clean, standardized, and adequately prepared for detailed analysis and visualization, forming a solid foundation for understanding the spread and evolution of COVID-19 variants across different regions and periods.

### Sliding Windows Implementation

The selection of time window for DPGR in a target region focus on dates of collection. Typically, the range of chosen time window ranges from 4 weeks to 12 weeks, considering both the good R^2^ fit and predefined P value threshold. For the region-specific analysis, the time windows are aligned with the periods when the target variants are predominant in a certain region of interest. For instance, in our continent-level analysis Omicron and Delta variants were predominant in each continent within the chosen time windows. Time windows are also influenced by the availability of data. The pairwise transmission fitness estimation analysis relies on having enough data points within each window to perform pairwise comparisons. For instance, the time window in Asia spans from December 2021 (Figure 3 C), reflecting the period when Omicron began to spread rapidly in that region. The length of the time windows varies slightly depending on the region due to the variances in first detection of a variant at a certain region of interest, however they are generally consistent to allow for comparative analysis across different regions. This consistency helps in comparing the slope values (indicating the transmission fitness) across regions.

To understand the significance of the estimated transmission fitness we considered a significance level of 0.05 and compared the p-value against this threshold to determine the statistical significance of the results. We also considered the R^2^ value or coefficient of determination while selecting the time windows to understand how well our regression model fits the observed data, indicating the proportion of the dependent variable (transmission fitness) can be predicted from the independent variable (time window). A higher R^2^ value helps to ensure that the suggested relationship between transmission fitness and the selected time window is strong and consistent. To facilitate more cross-regional comparison R^2^ value provides consistency, ensuring the time windows selected in the regions of interest are based on strong statistical relationships. To ensure data quality the high R^2^ value helps to filter out periods where the observed data is noisy. Both the R^2^ value and p-value help to ensure the observed relationships are statistically significant and the model prediction is strong and explanatory. Thus, incorporating the sliding windows, R^2^ value and p-value during the estimates we ensured that our estimated transmission fitness is statistically significant.

### Neighbor Join Tree

The neighbor-joining method is an agglomerative clustering algorithm to create the phylogenetic tree of species. This method does not consider the constant rate of evolution; hence the branch length from each node to the tips varies. It takes in input as genome sequence data and calculates the distance between each pair of taxa. The neighbor-joining algorithm uses the pairwise distance matrix to create an unresolved star-shaped tree that is iterated over a few steps to find the tree’s branch lengths. From the distance matrix, it calculates a Q-matrix and finds the distance between each pair of taxa. It joins the two taxa with a new node and connects with the central node. The Q-matrix is calculated using this formula:

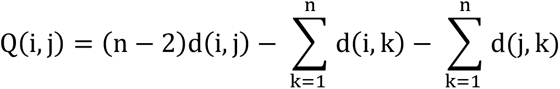

Then the algorithm calculates the distance of each of the taxa in the pair to the newly created node using the following equation:

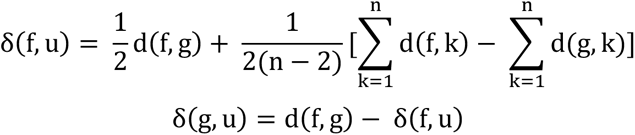

Here, u is the newly created node, and f, g is the taxa in the pair. The distance from the other taxa from the newly created node is calculated using the following equation:

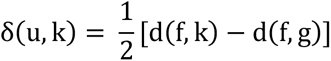

Here, k denotes the node to calculate the distance, and u refers to the newly created node.

### Distance Matrices

To construct the fitness stair and transmission fitness landscape, we created a pairwise distance matrix leveraging the Differential Population Growth Rate (DPGR) method to infer pairwise transmission fitness between SARS-CoV-2 variants. As the method is pairwise, we can directly estimate the transmission fitness of the adjacent variants like DPGR_Alpha,Beta_, DPGR_Beta, Delta_ and DPGR_Delta.Omicron_. To infer the transmission fitness of the non-adjacent variants, DPGR_Alpha,Delta_, DPGR_Alpha, Omicron_, the property of logarithms (log(a/b) = log(a/c) + log(c/b) is used. Thus, we can infer the growth advantages of non-adjacent variants. This technique allows us to create a distance matrix with a predefined root, capturing the relative fitness of multiple variants over a certain period. For our case, we considered Alpha, the earliest variant, as the root variant.

Here, the distance matrix is non-negative because we considered absolute pairwise fitness. In non-absolute relative terms, transmission fitness between Omicron to Alpha is negative and vice-versa.

### Construction of Fitness Stair

The fitness stair (See Figure 5 d) is a step plot generated using the plt.step() function from the Matplotlib library. This plot visually represents the progressive fitness growth of various COVID-19 Variants of Concern (VOCs) relative to a root variant, in this case, Alpha.

By examining the absolute pairwise distance matrix (as shown in Table 1), the fitness values for subsequent variants—Beta, Delta, and Omicron—progressively increase relative to Alpha. Specifically, the pairwise distances are 0.002313 for Beta, 0.008400 for Delta, and 0.017715 for Omicron. The distance between Alpha and itself is 0, as it serves as the reference point. This fitness stair (See Figure 5 d) effectively illustrates how each variant diverges in terms of fitness from the original Alpha variant, providing a clear visual representation of the evolutionary changes in their relative fitness.

**Table 1:**
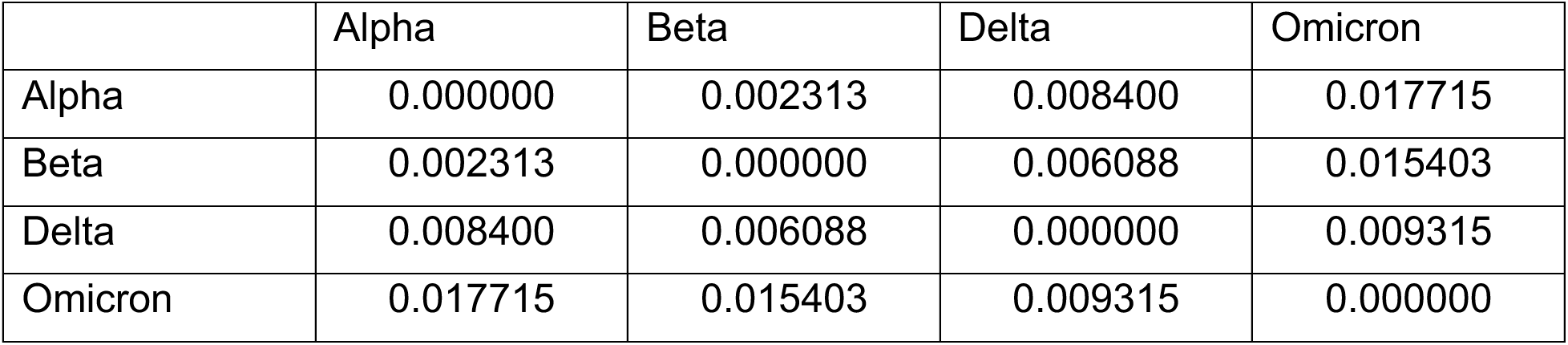
Pairwise Distance Matrix considering Alpha as the root variant.

|  | Alpha | Beta | Delta | Omicron |
| --- | --- | --- | --- | --- |
| Alpha | 0.000000 | 0.002313 | 0.008400 | 0.017715 |
| Beta | 0.002313 | 0.000000 | 0.006088 | 0.015403 |
| Delta | 0.008400 | 0.006088 | 0.000000 | 0.009315 |
| Omicron | 0.017715 | 0.015403 | 0.009315 | 0.000000 |

### Construction of Fitness Landscape

In evolutionary biology, reproductive success of the organisms is understood by the fitness landscape. In the fitness landscape, the height of the landscape defines the relative fitness of one genotype with respect to the others. In our study, using the pairwise absolute distance matrix (Table 1) we constructed a pairwise fitness landscape to understand the relative fitness of pairs of variants. The variant pairs on the top of the landscape have higher relative fitness than those on the bottom of the landscape.

The fitness landscape is generated by reading the distance matrix (Table 1) from a CSV file. This distance matrix, which contains pairwise fitness values between different SARS-CoV-2 variants, is used to create a 3D surface plot. The Python pandas library is utilized to load the data and set the appropriate columns as the DataFrame’s index. The x and y coordinates for the plot are derived from the matrix’s columns and index, respectively. Using Plotly’s “go.Surface” function, a surface trace is created, which visualizes the fitness values as a continuous 3D surface. The plot is configured to have transparent axes and background, ensuring a clean visualization focused on the fitness landscape itself. The camera position and perspective are adjusted to provide an optimal view of the surface to understand the relative fitness growth of the pairs of variants. To create the fitness landscape, we used the absolute pairwise distance matrix to visualize the landscape. As we go towards the peaks we observe the pairs with higher relative fitness. For instance, in Figure 5d, Omicron Vs. Alpha has higher relative fitness than Delta Vs. Omicron. This comprehensive visualization effectively highlights the relative fitness of each variant in a three-dimensional space, offering valuable insights into their evolutionary dynamics.

## Data Availability

The GISAID data can be accessed at https://gisaid.org/

## Acknowledgments

HQ thanks the NSF award PIPP #2200138, BD Spoke #1761839, REU #1852042 and #2149956, Tennessee AI Initiative, internal support of the Department of Computer Science and Engineering and UTC, and internal support of School of Data Science and the Department of Computer Science at Old Dominion University. SH is partially supported by NSF REU #1852042.

## Code and Data Availability

Sample code and important estimates are provided at https://github.com/QinLab/DPGR. The GISAID data can be accessed at https://gisaid.org/.

## Author Contributions

HQ conceived the idea and designed the research .MJP conducted the research, with assistance from LAB and SH .MJP and HQ drafted and revised the manuscript .ChatGPT 4.0 was used in polishing the languages of manuscript. All authors read and approved the final manuscript

## Supporting Document

### 1 Construction of the Differential Population Growth Rate (DPGR) Model

When estimating the Differential Population Growth Rate (DPGR) between two target variants, it is assumed that the populations exhibit exponential growth rates. A period of linear growth is selected for analysis within the time window between pairs of target variants. Additionally, a time lag is accounted for to address the difference in the timing of their growth phases.

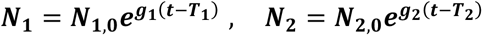

where, the variable t is time, N_1_ and N_2_ are the population as a function of time, N_1,0_ and N_2,0_ are the initial populations, g1 and g2 are the growth rates, and T_1_ and T_2_ are the time lag of their growth, where t > T_1_ and T_2_.

Now taking the ratio of their growth rate,

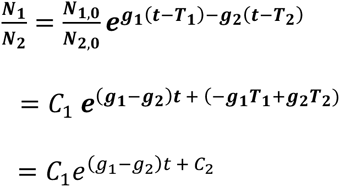

Here, C_1_ = N_1,0_/N_2,0_ and C_2_ = (-g_1_T_1_+ g_2_T_2_) are constants. Now taking the log transformation on both sides, we get,

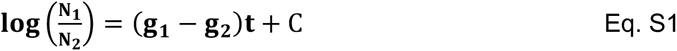

where C is a constant derived from C_1_ and C_2_. Because of the asymptotic cases of SAS-CoV-2, it is likely N_1_ and N_2_ are under-reported, which can lead to sampling biases. If we assume that the sampling biases are invariant or similar among different variants, then the ratio of N_1_/N_2_ would approximate the true ratio of the two subpopulations of variations.

The term (g_1_−g_2_) denotes the Differential Population between the two populations of interest. If g1 is greater than g2, the first population grows faster than the second, and DPGR is a positive value. Otherwise, DPGR would be a negative value.

With the linear form of Eq S1, we can apply linear regression to a scatter plot of log(N1/N2) versus t in an appropriate time window to infer DPGR.

### 2. Indirect Estimation of DPGR

During the COVID-19 pandemic period, some variants, say N_1_, dominated in earlier stages and some variants, say N_3_, appeared much later. Given the limited genomic sampling capacity, we cannot observe N_1_/N_3_ directly. In this case, we can choose an intermediate variant N2 through which we can observe N_1_/N_2_ and N_2_/N_3_ through genomic sampling. For instance, for SARS-CoV-2 variants, there is limited co-sampling of Alpha and Delta variants in the same location which makes DPGR_Alpha-Delta_ challenging to estimate. With the Beta variant as an intermediate, we can have DPGR_Alpha-Delta_ = DPGR _Alpha-Beta_ + DPGR _Beta-Delta_.

The inference is based on the property of logarithms indicates, log(a/c) =log(a/b)+log(b/c), and is illustrated as follows.

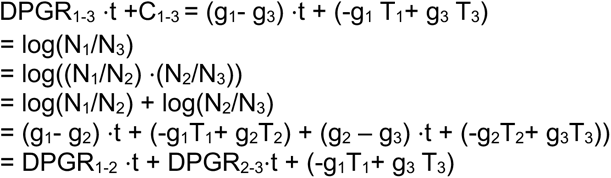

Simplifying, we have,

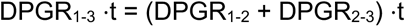

Hence,

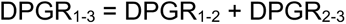

### 3 Mitigating the Sampling Biases in Genomic Surveillance Dataset and Validating Model Robustness against Noise

Existing literature have underscored the issue that estimating the viral transmission fitness is prone to sampling biases. Inconsistency in the data and geographic dominance of one variant over the other can highly compromise the estimated fitness values at a certain region^7^. Moreover, underreporting of the genomic sequencing data is frequently observed. For instance, in many of the African, Asian and South American countries underreporting is a crucial obstacle to estimating the viral transmission fitness. However, our proposed model made an effort to mitigate the sampling biases caused by an imbalanced dataset. One of the primary key points to be considered is the model estimates the pairwise transmission fitness. For estimating this pairwise transmission fitness, the weekly submission counts for any two target variants at each location within a particular time window are considered. This approach eliminates the chance of considering only the highly dominant variants in the target region and neglecting the less frequent ones. Thus, considering pairwise comparison and all the frequencies associated with target variants within the selected time window helps to reduce the biases induced by sampling, resulting in a more accurate estimation of transmission fitness, and strengthening the robustness of the model.

To test the model’s performance in response to the noise in the data, the preprocessed GISAID dataset is induced with random noise from the Gaussian Distribution, widely known as the Normal Distribution or Bell Curve. The noise is added to the ‘*Freq’* column of the dataset, which records the weekly sum of submission counts for each variant observed at a particular location. Gaussian distribution of mean 0 and varying standard deviation depending on the spread of the frequency count in a particular location is selected to induce the generated noise. Then the “np.normal.random()” function from the NumPy library is used to select random values from the Gaussian Distribution and added to the existing values of the *Freq* column. After that, the model is fitted with the noisy data to observe the linear performance of the model, as according to the assumption of the DPGR model, the transmission fitness growth must follow a linear pattern. Figure S7 and S8 illustrate the transmission fitness estimation scatterplots of Omicron compared to Delta in several countries and continents. It is apparent that after introducing the noise, the estimated fitness values changed, and the range of the maximum and minimum fitness shifted, which is expected; however, the observed growth in the pairwise transmission fitness of Omicron is still maintaining the linear trend with R^2^ values ranging from 0.864 to 0.995 for continent level estimation. Higher R^2^ values are observed in North America (0.974), Europe (0.995) and Oceania (0.985) and the lowest R^2^ value is observed in Asia (0.864) (details in Table S4). For the country level analysis of transmission fitness estimation under gaussian noise we observed R^2^ values ranging from 0.65 to 0.96 (details in Table S5). Therefore, the linearity of the model validates the model’s resiliency in handling the introduced noise, demonstrating the ability of the model to capture the variations.

Alongside adding random noise from the gaussian distribution, we also added synthetic noise to the target country (USA) from our existing distribution of Turkey to comprehend the robustness of the model by estimating the transmission fitness between Omicron and Delta between the sliding window of 2021-11-01 to 2021-12-31. We calculated the frequency of weekly submission counts for Turkey and added it to our target country, USA, for pairwise estimation. We mixed varying proportions (see details of the mix proportions in Table S3) of USA and Turkey submission counts to understand the noise handling performance of the model. For every combination of mix proportions the observed R^2^ value is 0.97. The purpose of this addition is to simulate an enhanced dataset with an induced variance, mirroring potential inconsistencies akin to real-world surveillance discrepancies.

Existing frequency count of USA, now supplemented with the frequency from Turkey, underwent pairwise transmission fitness estimation. The resultant transmission fitness and corresponding p-values (ranging from 0.00000194 to 0.00000189) offer nuanced insights into the model’s significant predictive stability and accuracy in the face of synthetically induced data variability. (details in Table S3)

This methodological fortification through synthetic noise introduction is crucial in our pursuit of model validation. It ensures that our model is not only calibrated to pristine datasets but can also withstand and accurately interpret datasets that have been subjected to the vicissitudes of real-world data collection and reporting anomalies.

### 4. Supporting Figures

**Figure S1:**
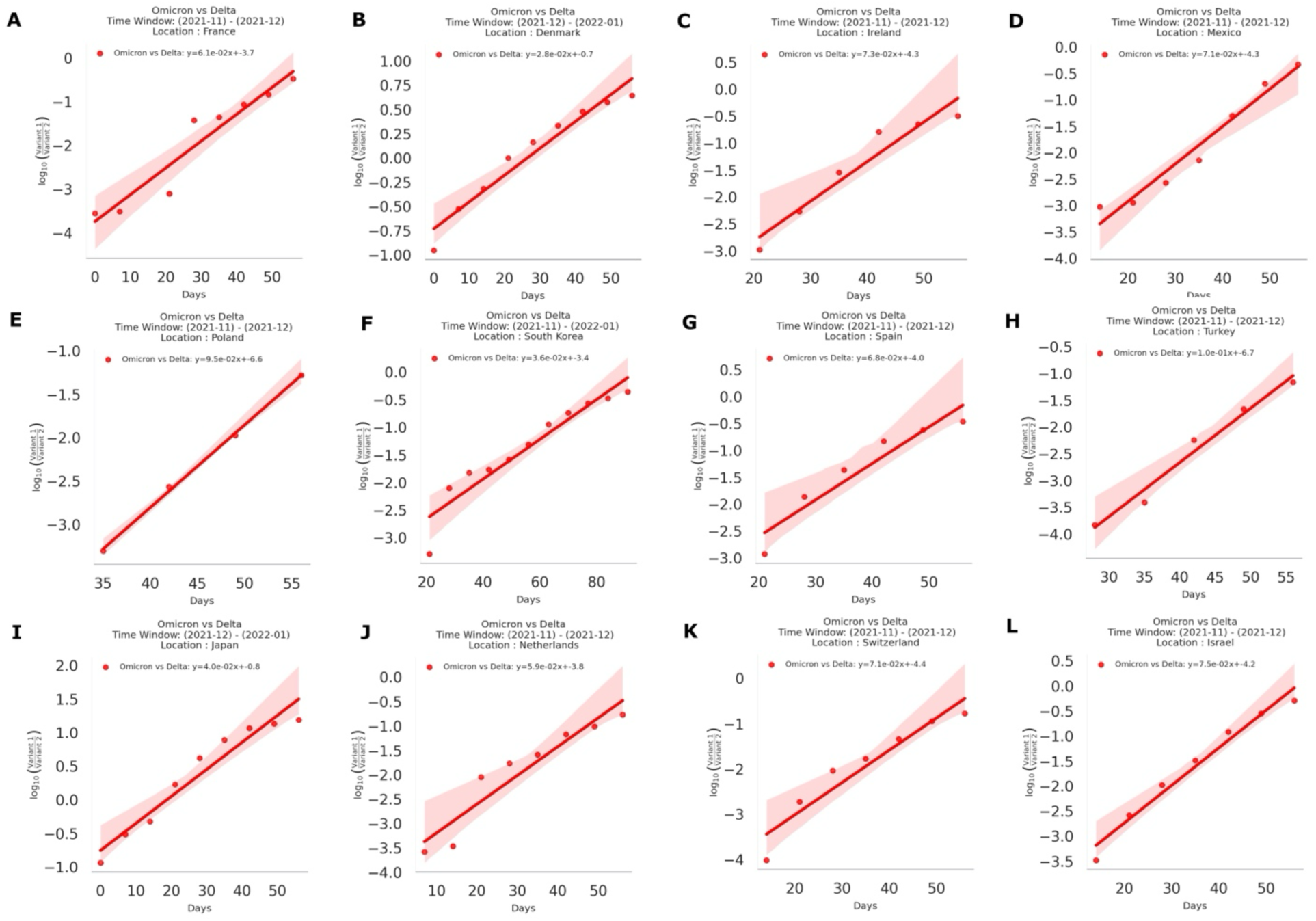
Pairwise Transmission fitness estimation of Omicron compared to Delta in several countries. The subplots (A-L) illustrate the sharp increase of pairwise transmission fitness of Omicron Compared to Delta in the target countries.

**Figure S2.**
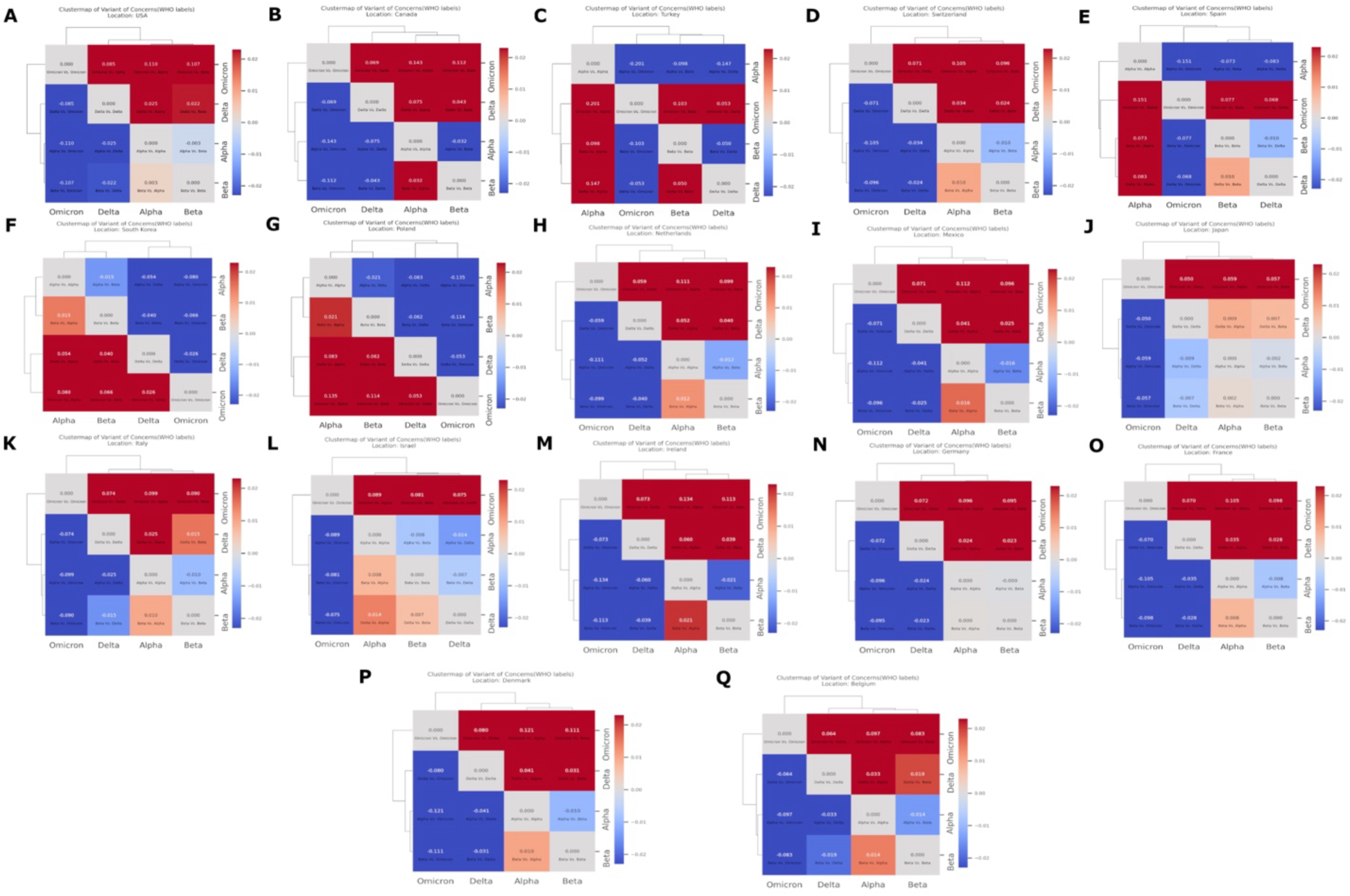
Heatmaps of the estimated pairwise transmission fitness of the WHO labeled Variants of Concern (VOCs) for the target countries. The heatmaps (A-Q) are read from row to column, describing the transmission fitness of a row variant compared to a variant in a column. The dark red cells in the figures indicate higher transmission fitness of the specific variant in the row compared to the variant in the column. Cells colored in blue indicate that a particular variant in the row has negative transmission fitness growth with the variant in the column. Dendrograms are plotted on the left and the top of each figure to depict differences in the transmission fitness growth rate of one variant to another. A higher variant distance indicates a significant difference in growth. Variants under the same node are closer to each other in transmission fitness growth than the other variants.

**Figure S3:**
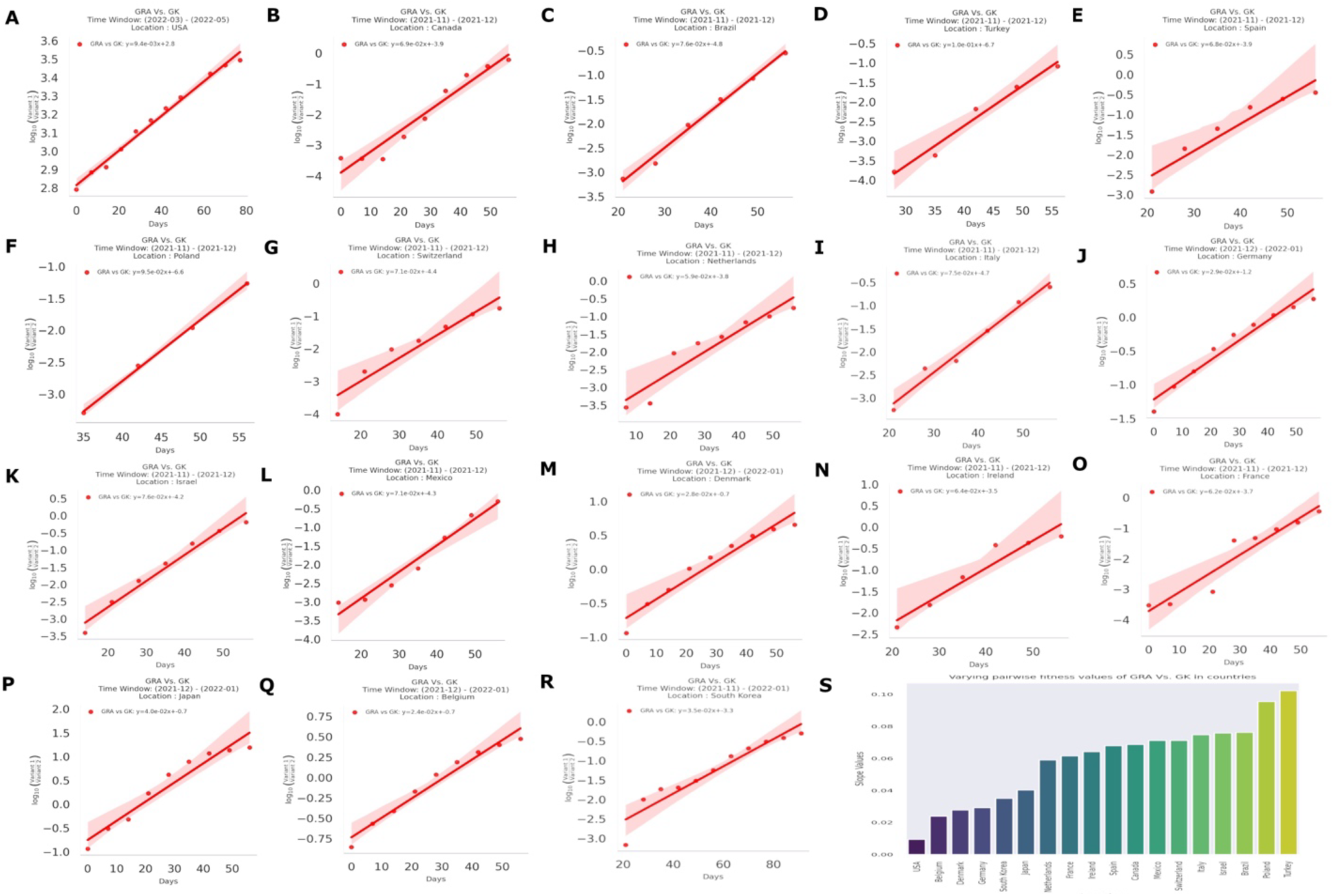
Pairwise Transmission fitness estimation of GRA compared to GK in target countries. The sub-plots **(A-G)** illustrate the estimated pairwise transmission fitness of GRA Compared to GK in the target countries. G. The bar plot visualizes the estimated transmission fitness values in all the analyzed regions.

**Figure S4:**
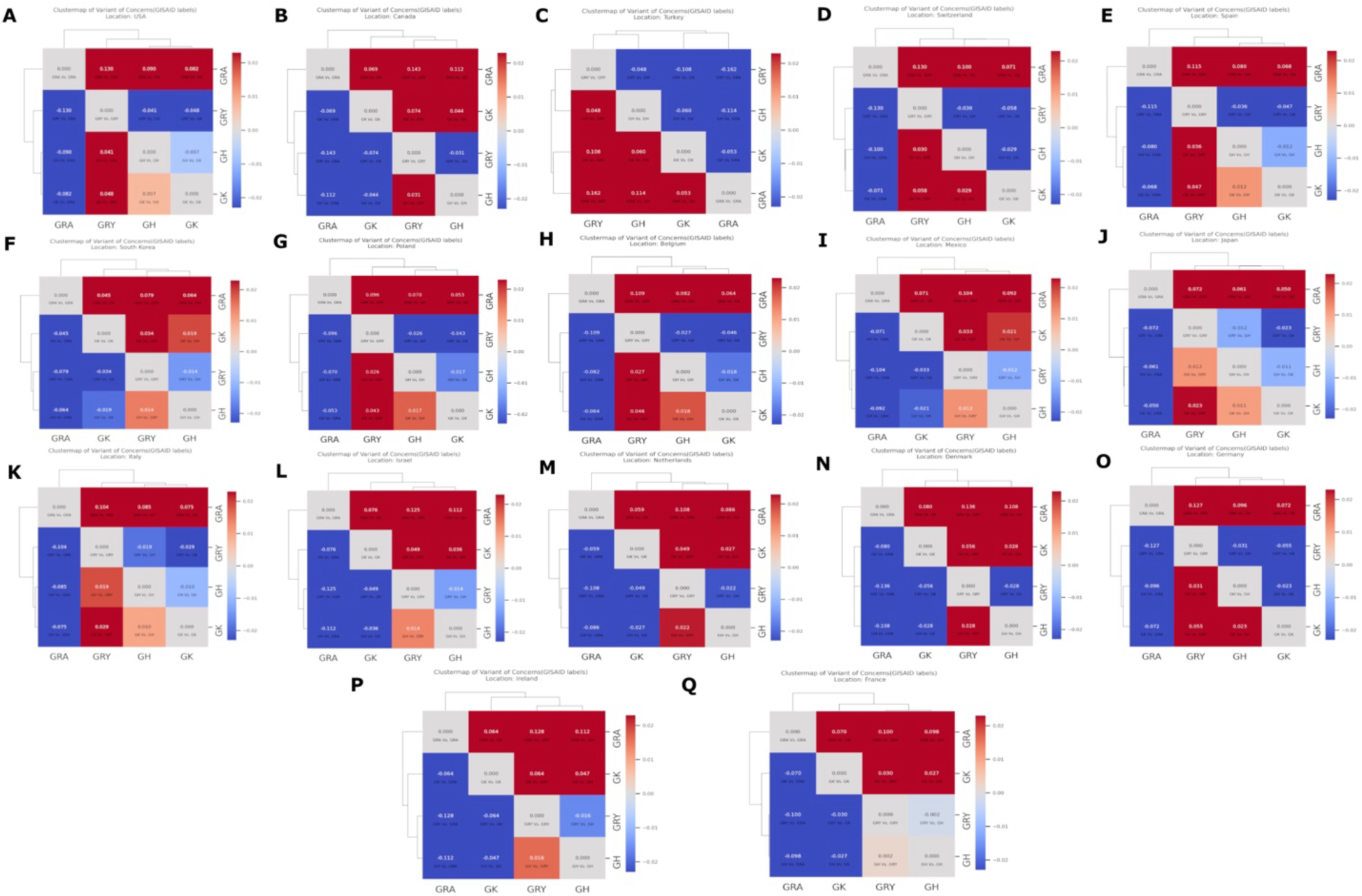
Heatmaps of the estimated pairwise transmission fitness of the GISAID labeled clades containing Variants of Concern (VOCs) for the target countries. The heatmaps from **(A-Q)** are read from row to column, describing the transmission fitness of a row variant compared to a variant in a column. The dark red cells in the figures indicate higher transmission fitness of the specific clade in the row compared to the clade in the column. Cells colored in blue indicate that a particular clade in the row has negative transmission fitness growth with the corresponding clade in the column. Dendrograms are plotted on the left and the top of each figure to depict differences in the transmission fitness growth rate of one clade to another. A higher clade distance indicates a significant difference in transmission fitness growth. Clades under the same node are closer to each other in transmission fitness growth than the other clades.

**Figure S5:**
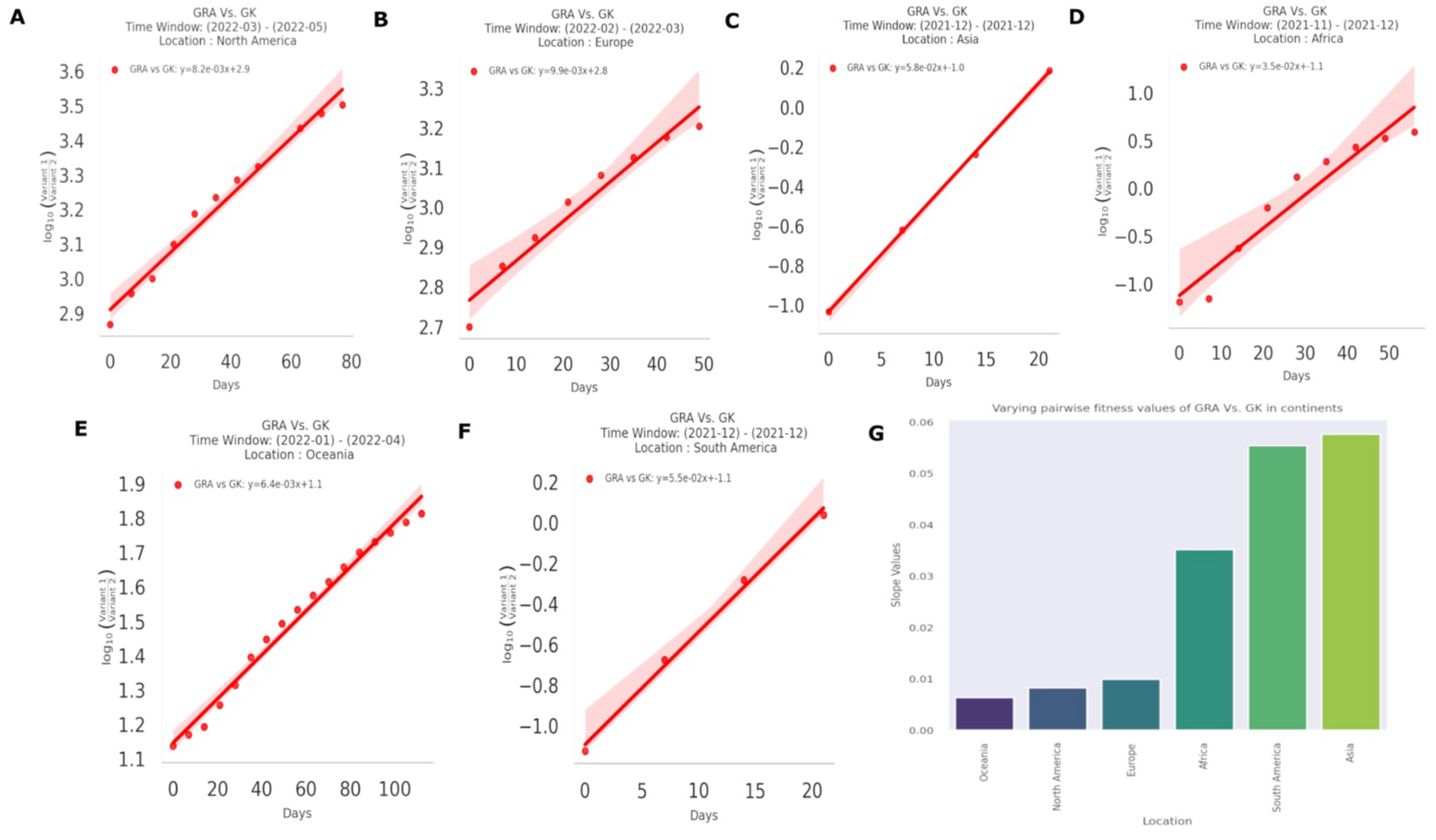
Pairwise Transmission fitness estimation of GRA compared to GK in target continents. The sub-plots **(A-F)** illustrate the sharp increase of pairwise transmission fitness of GRA Compared to GK in the target continents. **G.** The bar plot visualizes the estimated transmission fitness values in all the analyzed regions

**Figure S6:**
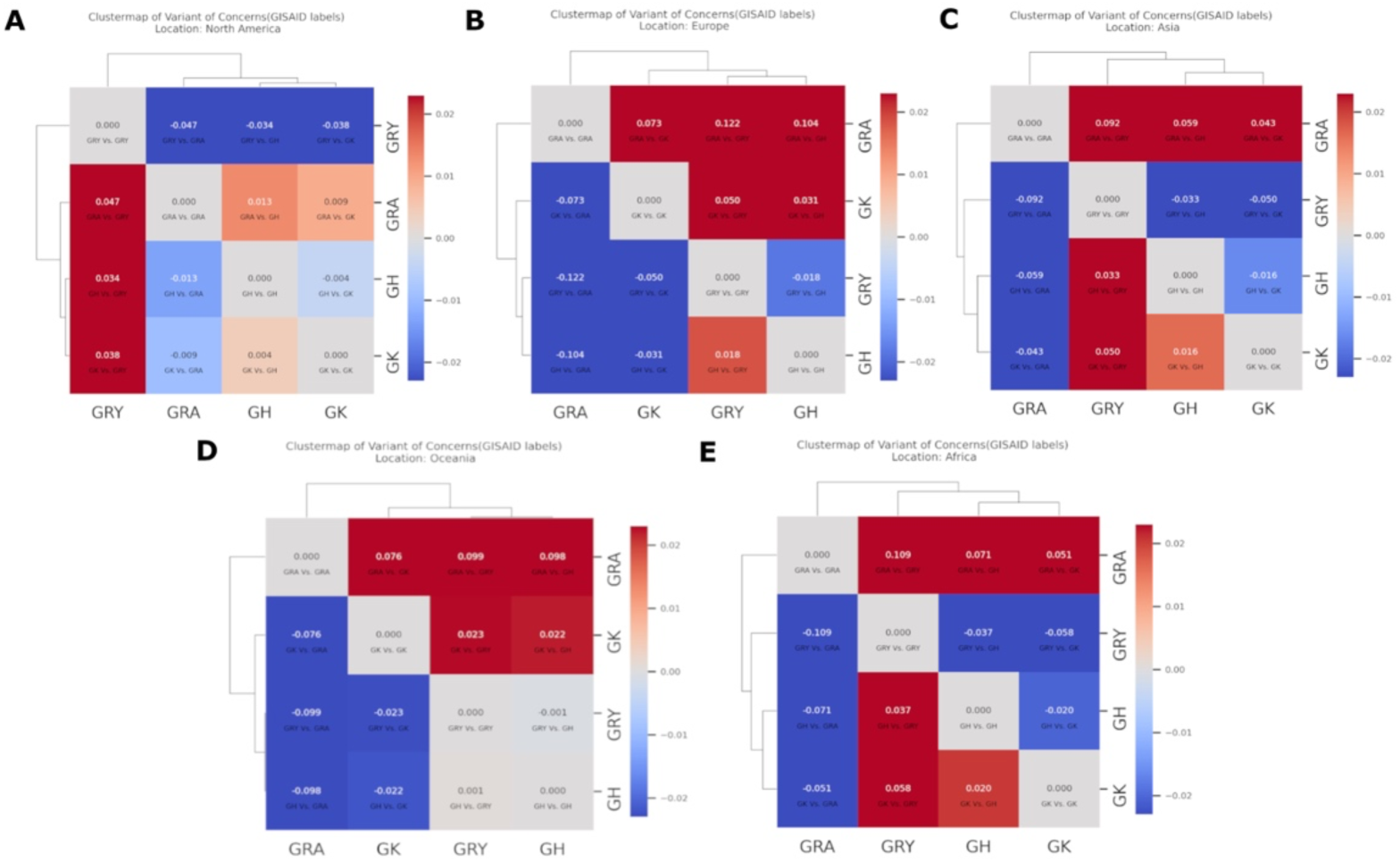
Heatmaps of the estimated pairwise transmission fitness of the GISAID labeled clades containing Variants of Concern (VOCs) for the target continents. The heatmaps from **(A-E)** are read from row to column, describing the transmission fitness of a row clade compared to a clade in a column. The dark red cells in the figures indicate higher transmission fitness of the specific clade in the row compared to the clade in the column. Cells colored in blue indicate that a particular clade in the row has negative transmission fitness growth with the corresponding clade in the column. Dendrograms are plotted on the left and the top of each figure to depict differences in the transmission fitness growth rate of one clade to another. A higher clade distance indicates a significant difference in transmission fitness growth. Clades under the same node are closer to each other in transmission fitness growth than the other clades.

**Figure S7:**
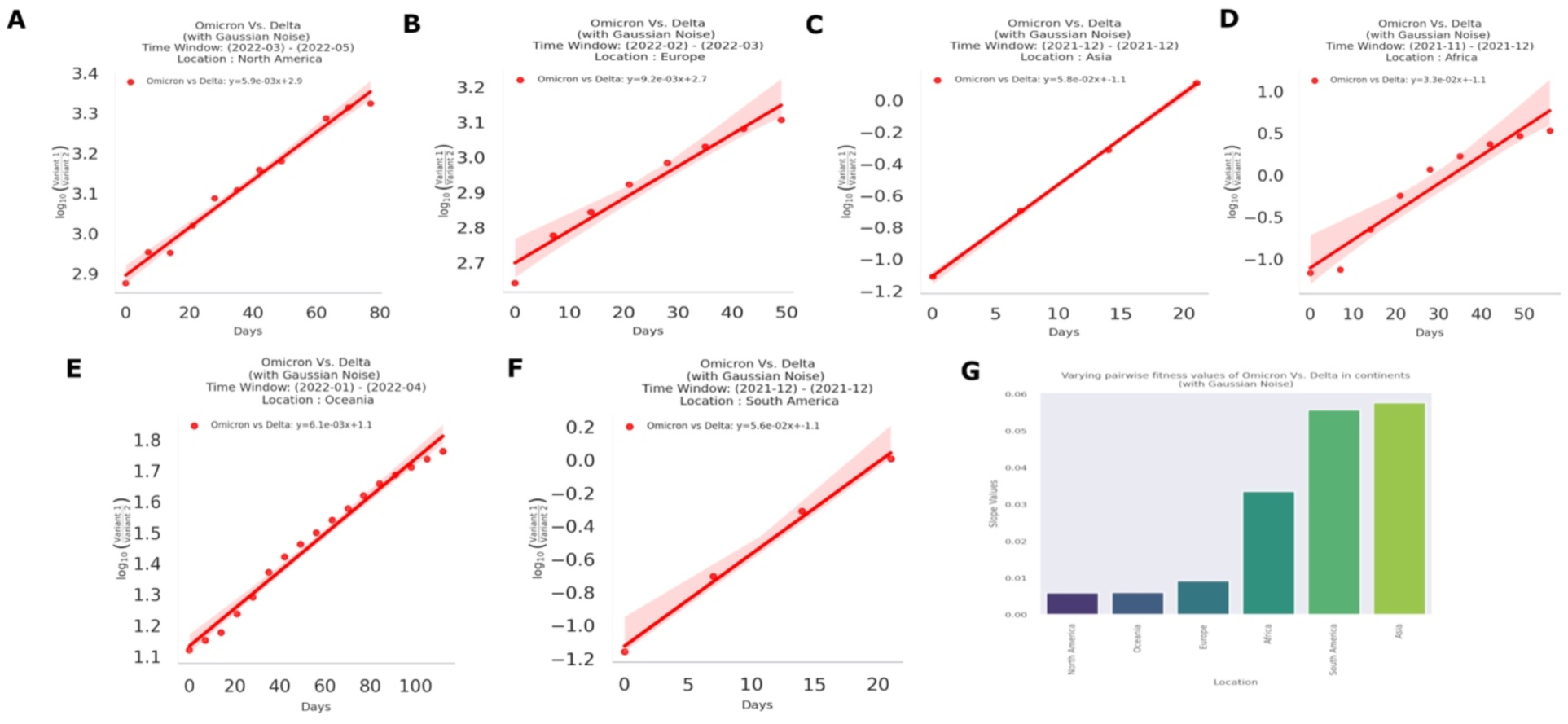
Pairwise Transmission fitness estimation of Omicron compared to Delta after introducing Gaussian noise in target continents. The estimated transmission fitness plots **(A-R)** of Omicron Compared to Delta in the target countries after introducing Gaussian Noise to validate model performance under simulated noises. The bar plot visualizes the estimated transmission fitness values in all the analyzed regions.

**Figure S8:**
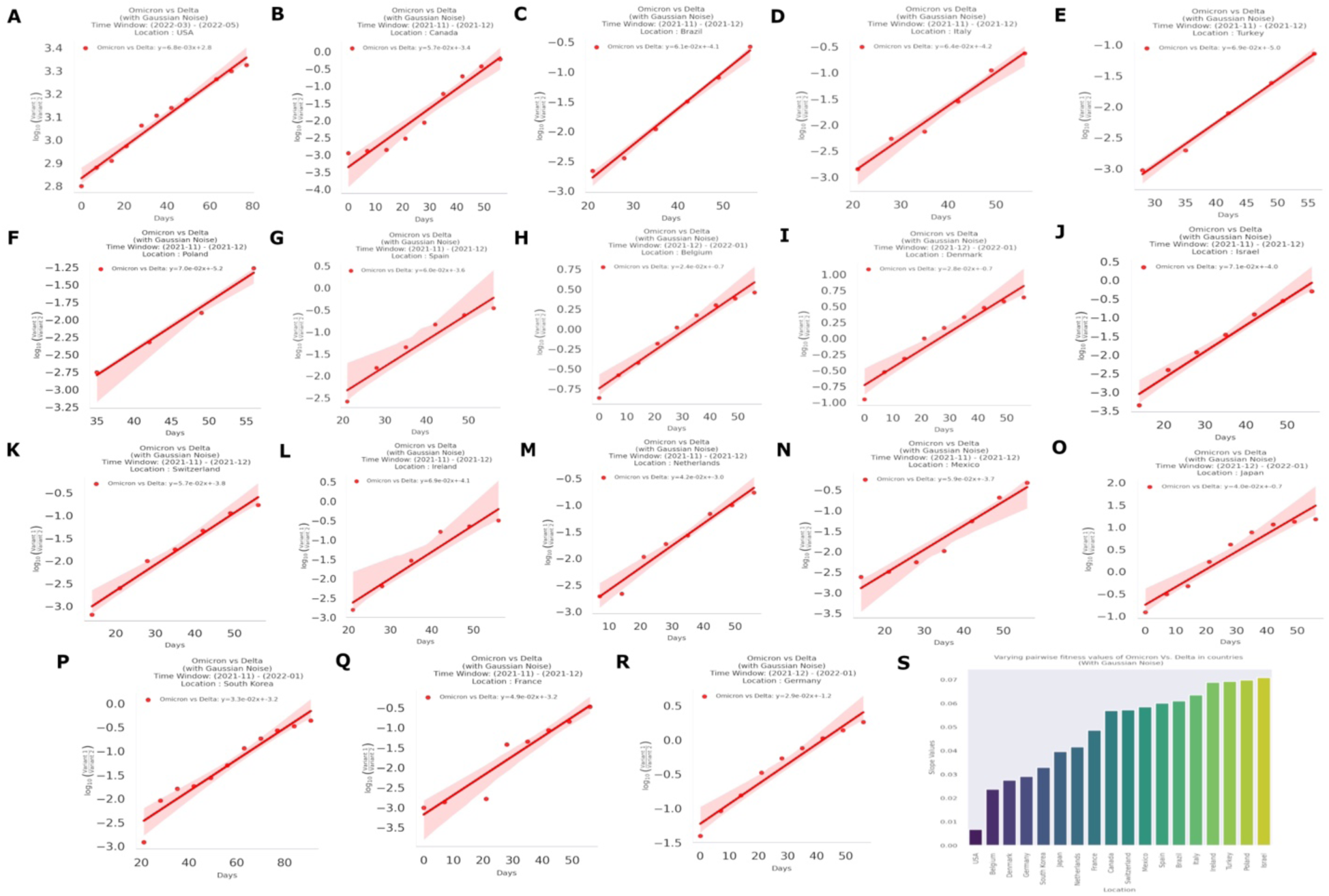
Pairwise Transmission fitness estimation of Omicron compared to Delta after introducing Gaussian noise in target countries. The estimated transmission fitness plots **(A-F)** of Omicron Compared to Delta in the target different continents after introducing Gaussian Noise. **G**. Bar plot visualizes the estimated transmission fitness values in all the analyzed regions.

**Figure S9:**
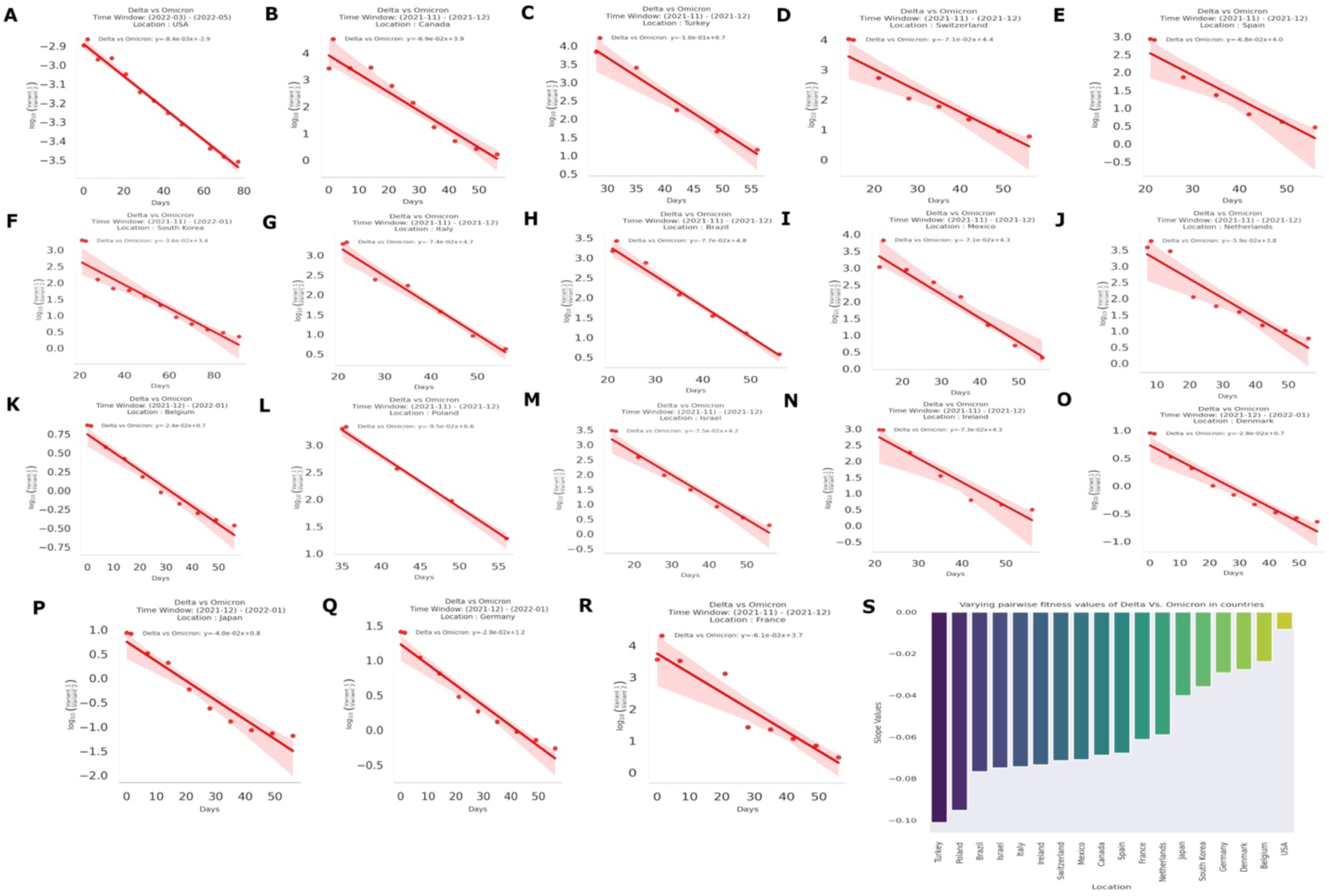
Estimated pairwise transmission fitness of Delta Compared to Omicron in several countries. The sub-plots **(A-R)** illustrate transmission fitness plots of Delta Compared to Omicron. The negative trend line indicates Delta has a negative transmission fitness growth compared to Omicron in different time windows in the selected regions. **G.** Bar plot visualizes the estimated transmission fitness values in all the analyzed regions.

**Figure S10:**
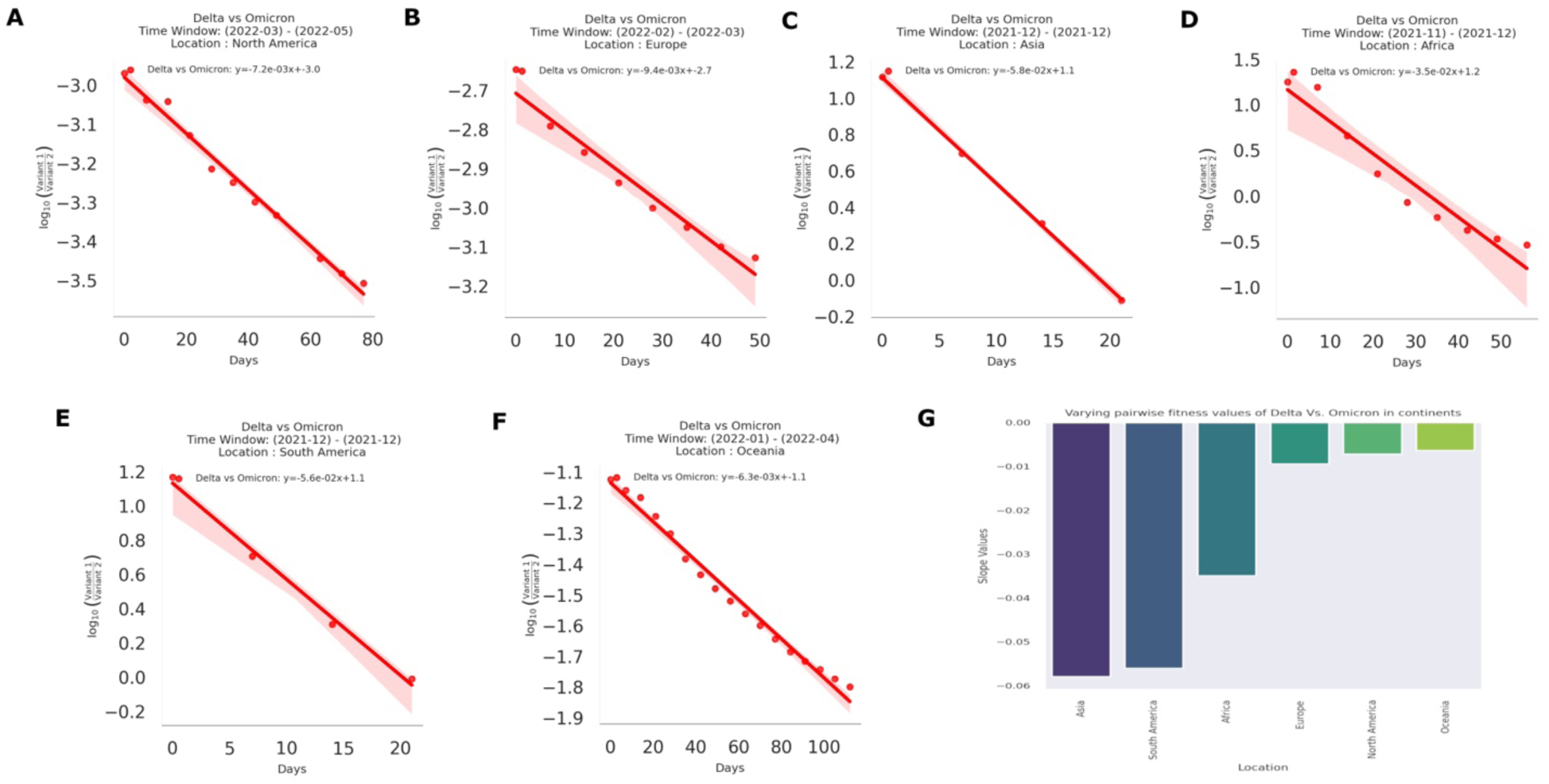
Estimated pairwise transmission fitness of Delta Compared to Omicron in several continents. The sub-plots (A-G) illustrate transmission fitness plots of Delta Compared to Omicron in the target continents. The negative trend line indicates Delta has a negative transmission fitness growth compared to Omicron in different time windows in the selected regions. G. Bar plot visualizes the estimated transmission fitness values in all the analyzed regions.

**Figure S11:**
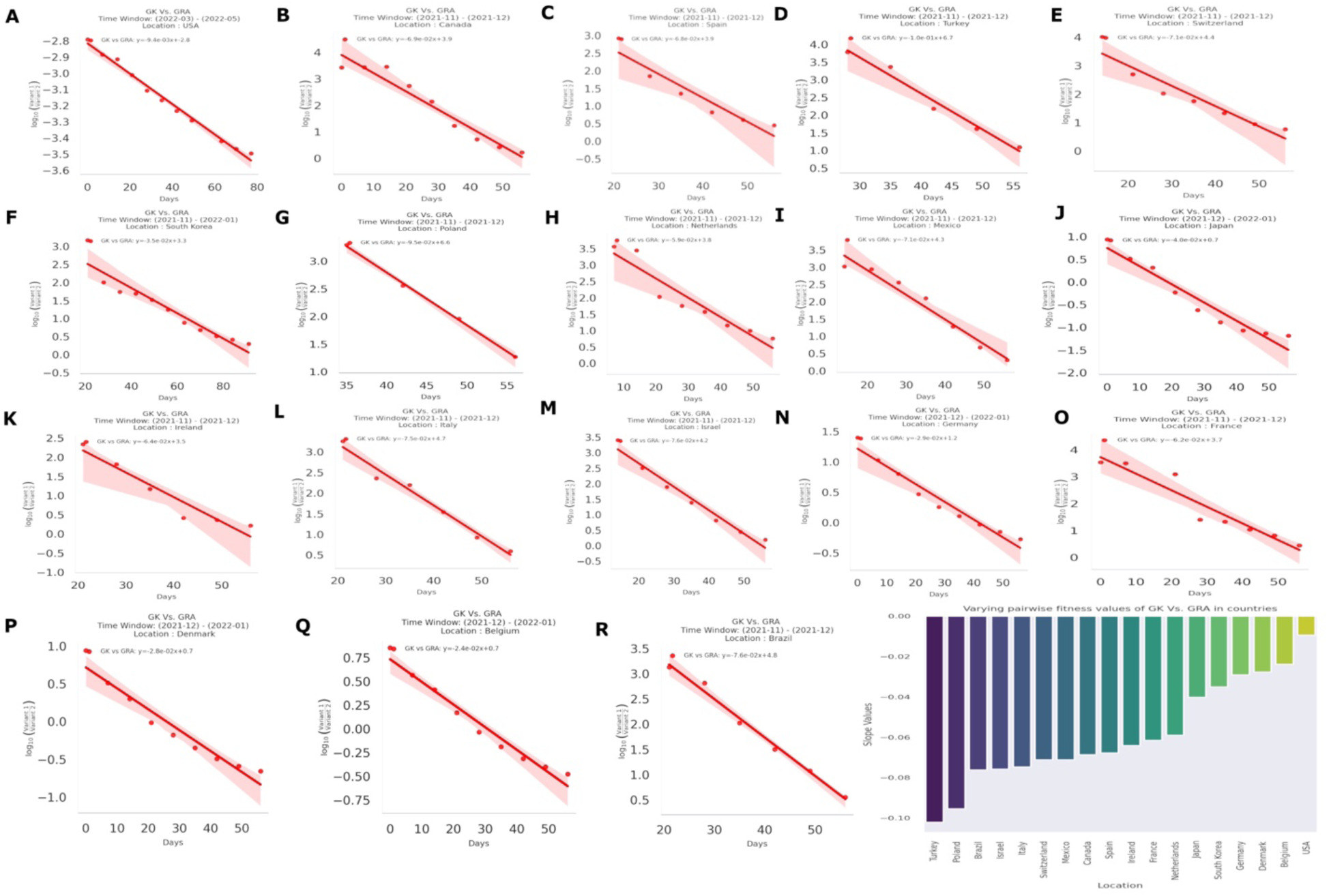
Estimated pairwise transmission fitness of GK Compared to GRA in several countries. The sub-plots **(A-R)** illustrate transmission fitness plots of GISAID clades GK Compared to GRA in the target countries. The negative trend line indicates GK has a negative transmission fitness growth compared to GRA in different time windows in the selected regions. **The** Bar plot visualizes the estimated transmission fitness values in all the analyzed regions.

**Figure S12:**
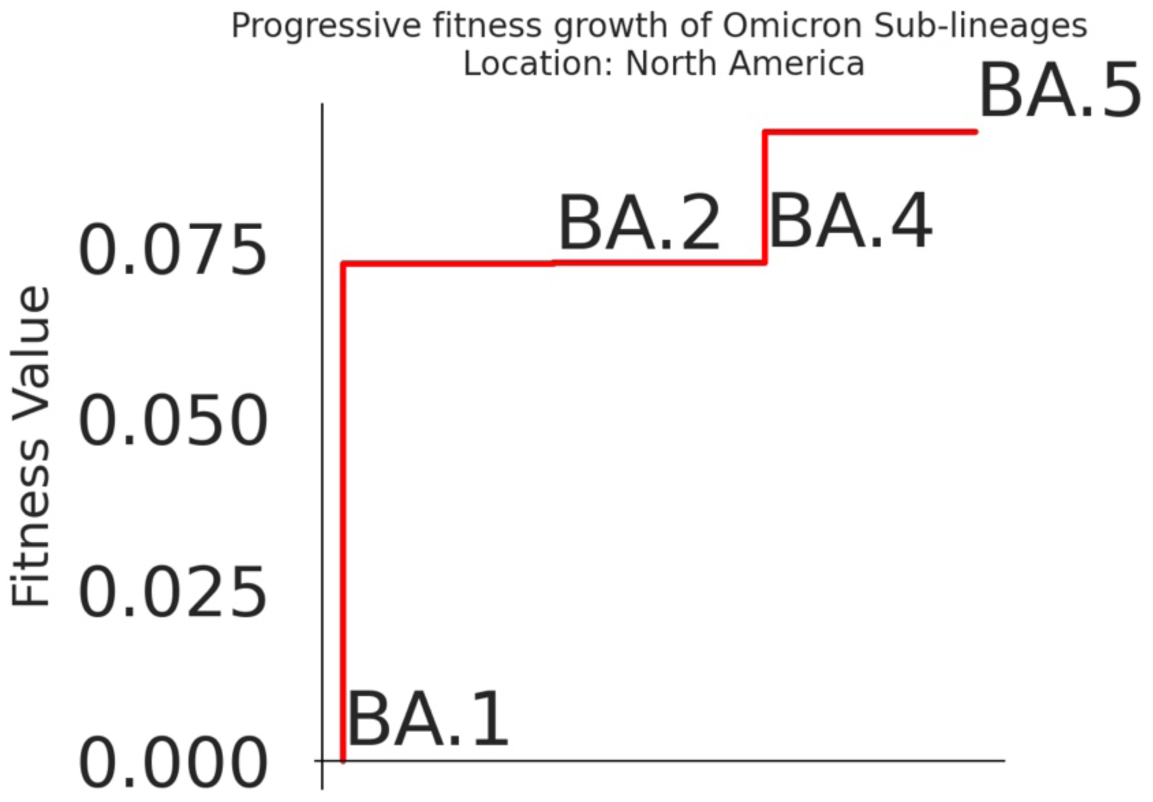
Fitness stair of Omicron Sub-lineages (BA.1*, BA.2*, BA.4*, BA.5*) The fitness stair illustrates the progressive transmission fitness gain of the Sub-lineages of the Omicron variant in the United States.

**Figure S13:**
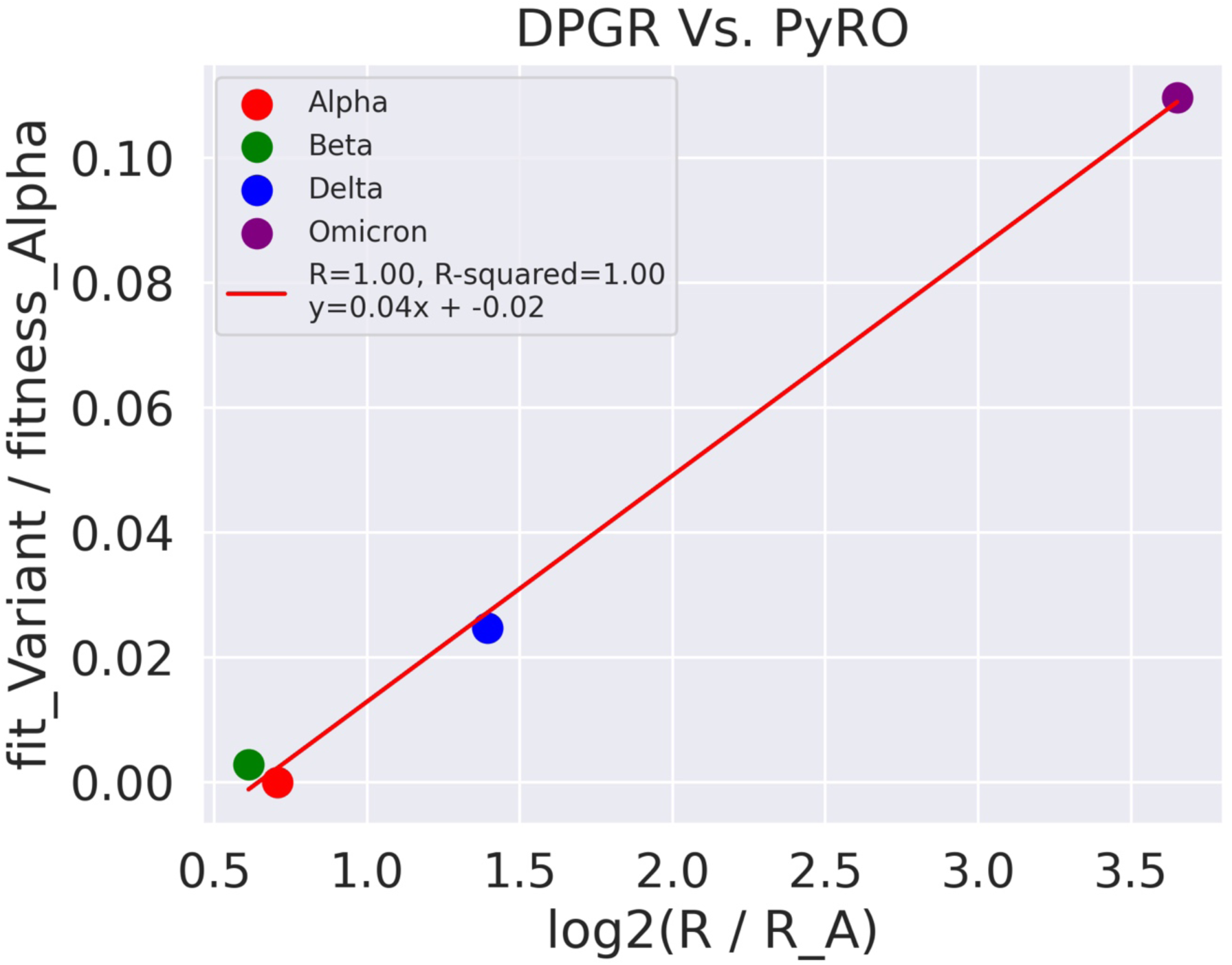
Estimated transmission fitness comparison between DPGR and PyRO model.

**Figure S14:**
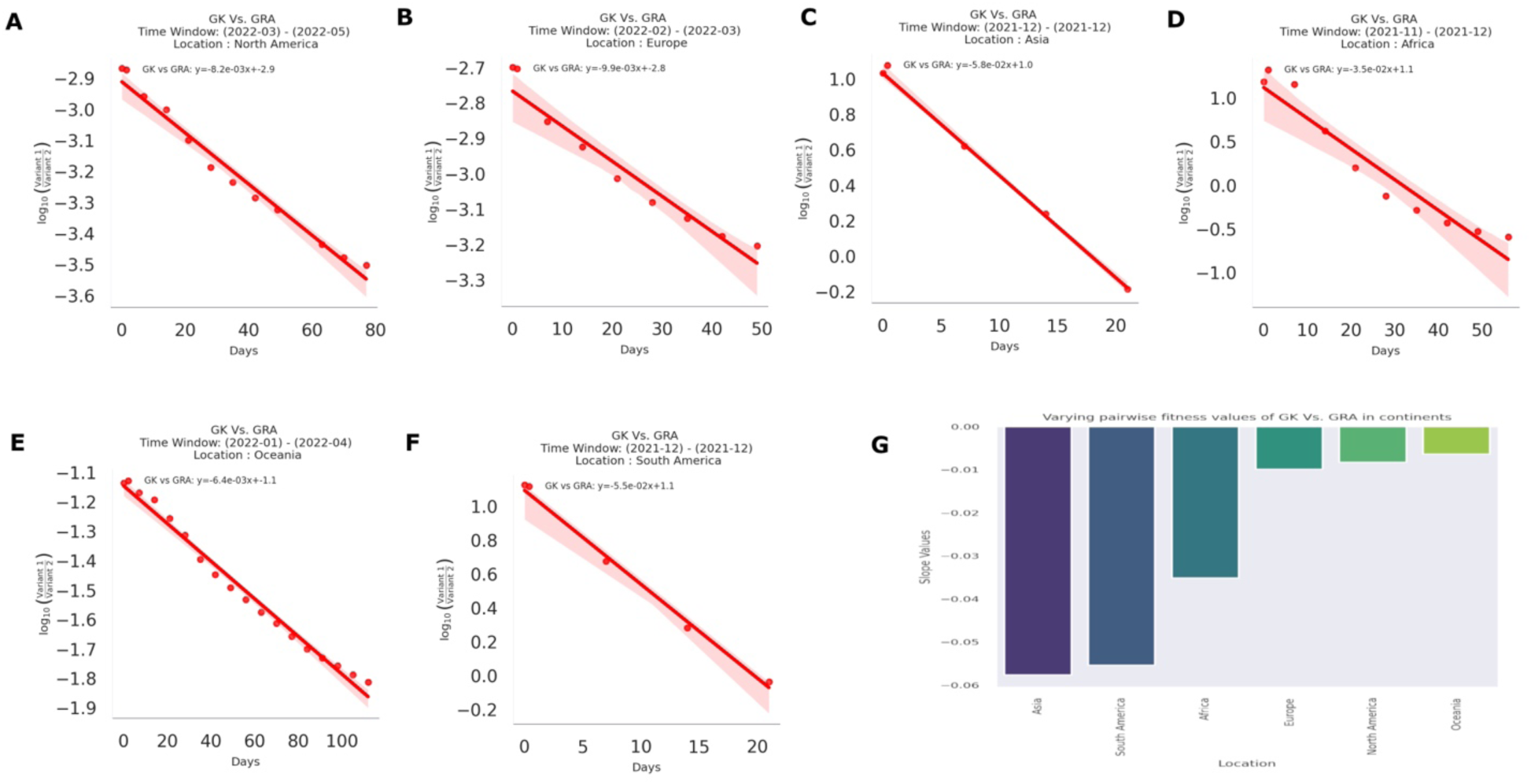
Estimated pairwise transmission fitness of GK Compared to GRA in several continents. The sub-plots **(A-F)** illustrate transmission fitness plots of GISAID clades GK Compared to GRA in the target continents. The negative trend line indicates GK has a negative transmission fitness growth compared to GRA in different time windows in the selected regions. **G.** Bar plot visualizes the estimated transmission fitness values in all the analyzed regions.

### 5. Supporting Tables

**Table S1:** Observed R^2^ Values for Different Geographic Locations for Omicron Vs. Delta:

| Location (Country) | P-Value | Slope Value | $R^2$ value | Time Window |
| --- | --- | --- | --- | --- |
| USA | 0.0000000002 | 0.0084 | 0.99 | '2022-03' - '2022-05' |
| Canada | 0.0000121512 | 0.0688 | 0.94 | '2021-11' - '2021-12' |
| Brazil | 0.0000424733 | 0.0767 | 0.98 | '2021-11' - '2021-12' |
| Germany | 0.0000037926 | 0.0292 | 0.96 | '2021-12' - '2022-01' |
| Belgium | 0.0000017600 | 0.0239 | 0.97 | '2021-12' - '2022-01' |
| Italy | 0.0002011518 | 0.0743 | 0.98 | '2021-11' - '2021-12' |
| Turkey | 0.0017071813 | 0.1011 | 0.97 | '2021-11' - '2021-12' |
| Israel | 0.0000498947 | 0.0750 | 0.97 | '2021-11' - '2021-12' |
| Ireland | 0.0019830378 | 0.0734 | 0.93 | '2021-11' - '2021-12' |
| Spain | 0.0034588120 | 0.0680 | 0.91 | '2021-11' - '2021-12' |
| France | 0.0002099997 | 0.0612 | 0.91 | '2021-11' - '2021-12' |
| Denmark | 0.0000116662 | 0.0280 | 0.94 | '2021-12' - '2022-01' |
| South Korea | 0.0000074797 | 0.0360 | 0.90 | '2021-11' - '2022-01' |
| Japan | 0.0000169371 | 0.0402 | 0.94 | '2021-12' - '2022-01' |
| Netherlands | 0.0003426374 | 0.0590 | 0.898 | '2021-11' - '2021-12' |
| Switzerland | 0.0008614151 | 0.0714 | 0.91 | '2021-11' - '2021-12' |
| Poland | 0.0007941103 | 0.0952 | 0.99 | '2021-11' - '2021-12' |
| Mexico | 0.0001007758 | 0.0709 | 0.96 | '2021-11' - '2021-12' |

| Location (Continents) | P-Value | Slope Value | $R^2$ value | Time Window |
| --- | --- | --- | --- | --- |
| North America | 0.0000000006 | 0.0072 | 0.99 | '2022-03' - '2022-05' |
| South America | 0.0032756997 | 0.0561 | 0.99 | '2021-12' |
| Europe | 0.0000234708 | 0.0094 | 0.96 | '2022-02' - '2022-03' |
| Oceania | 0.0000000000 | 0.0063 | 0.99 | '2022-01', '2022-02', '2022-03', '2022-04' |
| Asia | 0.0001804061 | 0.0579 | 0.98 | '2021-12' |
| Africa | 0.0000349938 | 0.0349 | 0.92 | '2021-11' - '2021-12' |

**Table S2:** Observed R^2^ Values for Different Geographic Locations for Omicron Sub-lineages (BA.1* -BA.5*)

| Location (Continent) | Comparison | P-Value | Slope Value | R<sup>2</sup> value | Time Window |
| --- | --- | --- | --- | --- | --- |
| North America | BA.5 vs BA.1 | 0.0000017075 | 0.053027 | 0.982 | '2022-04' – '2022-05' |
| North America | BA.5 vs BA.2 | 0.0000008811 | 0.042768 | 0.985 | '2022-04' – '2022-05' |
| North America | BA.5 vs BA.3 | 0.0002200994 | 0.052902 | 0.993 | '2022-04' – '2022-05' |
| North America | BA.5 vs BA.4 | 0.0001053811 | 0.013199 | 0.931 | '2022-04' – '2022-05' |
| Europe | BA.5 vs BA.1 | 0.0000000001 | 0.045530 | 0.986 | '2022-02' – '2022-05' |
| Europe | BA.5 vs BA.2 | 0.0000000001 | 0.042186 | 0.988 | '2022-02' – '2022-05' |
| Europe | BA.5 vs BA.3 | 0.0000000019 | 0.047117 | 0.990 | '2022-02' – '2022-05' |
| Europe | BA.5 vs BA.4 | 0.0047140545 | 0.003869 | 0.567 | '2022-02' – '2022-05' |
| Asia | BA.5 vs BA.1 | 0.0000056861 | 0.057299 | 0.974 | '2022-03' – '2022-05' |
| Asia | BA.5 vs BA.2 | 0.0000041944 | 0.046805 | 0.959 | '2022-03' – '2022-05' |
| Asia | BA.5 vs BA.3 | 0.0024389686 | 0.051755 | 0.968 | '2022-03' – '2022-05' |
| Asia | BA.5 vs BA.4 | 0.2765494733 | 0.005945 | 0.193 | '2022-03' – '2022-05' |
| Africa | BA.5 vs BA.1 | 0.0000000005 | 0.033623 | 0.988 | '2022-02' – '2022-04' |
| Africa | BA.5 vs BA.2 | 0.0000000042 | 0.027082 | 0.981 | '2022-02' – '2022-04' |
| Africa | BA.5 vs BA.3 | 0.0002743433 | 0.033385 | 0.993 | '2022-02' – '2022-04' |
| Oceania | BA.5 vs BA.1 | 0.0000000300 | 0.048157 | 0.995 | '2022-03' – '2022-05' |
| Oceania | BA.5 vs BA.2 | 0.0000000255 | 0.043159 | 0.995 | '2022-03' – '2022-05' |

**Table S3:**
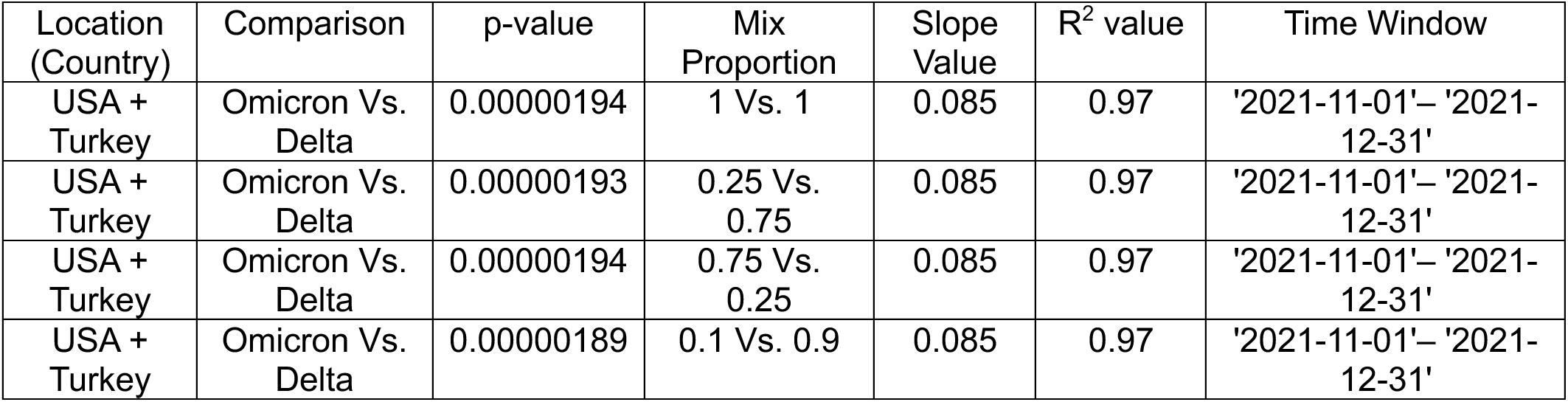
Observed R^2^ Values for Estimation by Introducing Synthetic Bias by Mixing Two Different Locations:

| Location (Country) | Comparison | p-value | Mix Proportion | Slope Value | $R^2$ value | Time Window |
| --- | --- | --- | --- | --- | --- | --- |
| USA + Turkey | Omicron Vs. Delta | 0.00000194 | 1 Vs. 1 | 0.085 | 0.97 | '2021-11-01'– '2021-12-31' |
| USA + Turkey | Omicron Vs. Delta | 0.00000193 | 0.25 Vs. 0.75 | 0.085 | 0.97 | '2021-11-01'– '2021-12-31' |
| USA + Turkey | Omicron Vs. Delta | 0.00000194 | 0.75 Vs. 0.25 | 0.085 | 0.97 | '2021-11-01'– '2021-12-31' |
| USA + Turkey | Omicron Vs. Delta | 0.00000189 | 0.1 Vs. 0.9 | 0.085 | 0.97 | '2021-11-01'– '2021-12-31' |

**Table S4:**
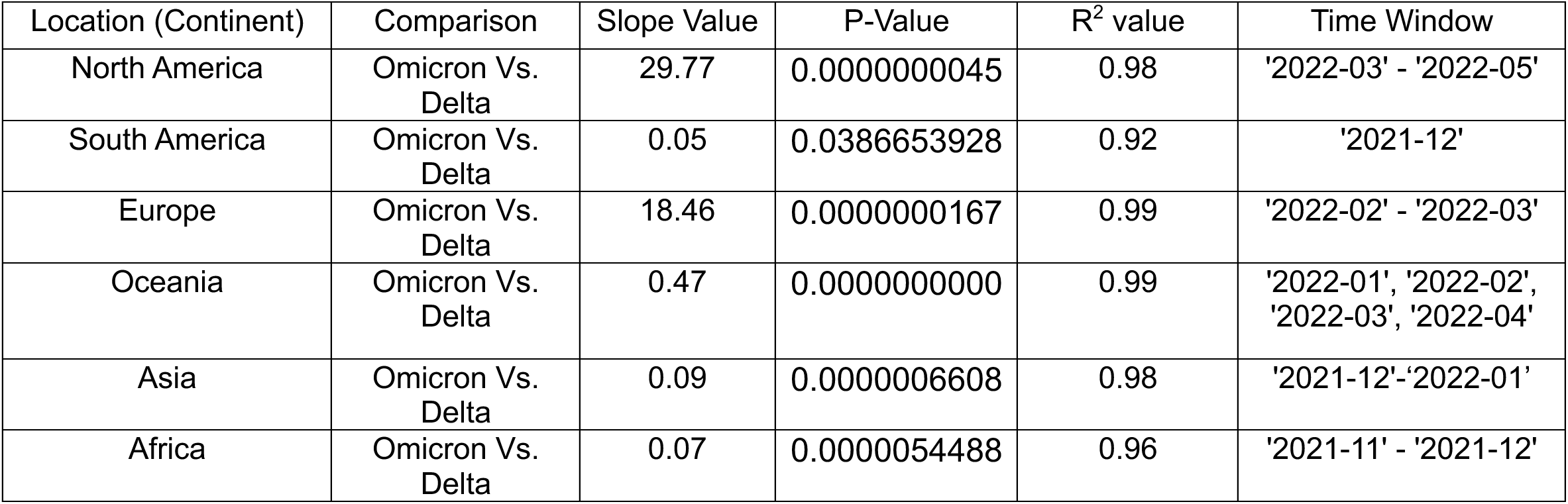
Observed R^2^ Values for Estimation by Introducing Gaussian Noise:

**Table S5:** Observed R^2^ Values for Estimation by Introducing Gaussian Noise:

| Location (Country) | Comparison | P-Value | Slope Value | $R^2$ value | Time Window |
| --- | --- | --- | --- | --- | --- |
| USA | Omicron Vs. Delta | 0.0000000981 | 35.67 | 0.96 | '2022-03' - '2022-05' |
| Canada | Omicron Vs. Delta | 0.0046773058 | 0.009 | 0.70 | '2021-11' - '2021-12' |
| Brazil | Omicron Vs. Delta | 0.0463248792 | 0.006 | 0.67 | '2021-11' - '2021-12' |
| Germany | Omicron Vs. Delta | 0.0000215862 | 0.031 | 0.93 | '2021-12' - '2022-01' |
| Belgium | Omicron Vs. Delta | 0.0000030266 | 0.051 | 0.96 | '2021-12' - '2022-01' |
| Italy | Omicron Vs. Delta | 0.0268002067 | 0.006 | 0.75 | '2021-11' - '2021-12' |
| Turkey | Omicron Vs. Delta | 0.0818599314 | 0.003 | 0.84 | '2021-11' - '2021-12' |
| Israel | Omicron Vs. Delta | 0.0104577546 | 0.014 | 0.84 | '2021-11' - '2021-12' |
| Ireland | Omicron Vs. Delta | 0.0024074943 | 0.009 | 0.92 | '2021-11' - '2021-12' |
| Spain | Omicron Vs. Delta | 0.0019413457 | 0.010 | 0.93 | '2021-11' - '2021-12' |
| France | Omicron Vs. Delta | 0.0146589756 | 0.005 | 0.66 | '2021-11' - '2021-12' |
| Denmark | Omicron Vs. Delta | 0.0000037399 | 0.081 | 0.96 | '2021-12' - '2022-01' |
| South Korea | Omicron Vs. Delta | 0.0000567553 | 0.006 | 0.85 | '2021-11' - '2022-01' |
| Japan | Omicron Vs. Delta | 0.0000279855 | 0.31 | 0.93 | '2021-12' - '2022-01' |
| Netherlands | Omicron Vs. Delta | 0.0027182864 | 0.003 | 0.86 | '2021-11' - '2021-12' |
| Switzerland | Omicron Vs. Delta | 0.0055076478 | 0.004 | 0.81 | '2021-11' - '2021-12' |
| Poland | Omicron Vs. Delta | 0.1311799352 | 0.002 | 0.76 | '2021-11' - '2021-12' |
| Mexico | Omicron Vs. Delta | 0.0263814437 | 0.009 | 0.66 | '2021-11' - '2021-12' |

**Table S6:** Observed R^2^ Values for Different Geographic Locations for GRA Vs. GK:

| Location (Country) | P-Value | Slope Value | $R^2$ value | Time Window |
| --- | --- | --- | --- | --- |
| USA | 0.0000000003 | 0.0094 | 0.99 | '2022-03' - '2022-05' |
| Canada | 0.0000110765 | 0.0687 | 0.95 | '2021-11' - '2021-12' |
| Brazil | 0.0000332042 | 0.0762 | 0.99 | '2021-11' - '2021-12' |
| Germany | 0.0000038165 | 0.0292 | 0.96 | '2021-12' - '2022-01' |
| Belgium | 0.0000017206 | 0.0239 | 0.97 | '2021-12' - '2022-01' |
| Italy | 0.0001901356 | 0.0747 | 0.98 | '2021-11' - '2021-12' |
| Turkey | 0.0017446336 | 0.1021 | 0.97 | '2021-11' - '2021-12' |
| Israel | 0.0000478457 | 0.0758 | 0.97 | '2021-11' - '2021-12' |
| Ireland | 0.0023451291 | 0.0641 | 0.92 | '2021-11' - '2021-12' |
| Spain | 0.0035146910 | 0.0678 | 0.90 | '2021-11' - '2021-12' |
| France | 0.0002081598 | 0.0615 | 0.91 | '2021-11' - '2021-12' |
| Denmark | 0.0000116616 | 0.0277 | 0.94 | '2021-12' - '2022-01' |
| South Korea | 0.0000069824 | 0.0351 | 0.90 | '2021-11' - '2022-01' |
| Japan | 0.0000169799 | 0.0402 | 0.94 | '2021-12' - '2022-01' |
| Netherlands | 0.0003454587 | 0.0590 | 0.89 | '2021-11' - '2021-12' |
| Switzerland | 0.0009175994 | 0.0711 | 0.91 | '2021-11' - '2021-12' |
| Poland | 0.0007546938 | 0.0954 | 0.99 | '2021-11' - '2021-12' |
| Mexico | 0.0000858866 | 0.0711 | 0.96 | '2021-11' - '2021-12' |

| Location (Continents) | P-Value | Slope Value | $R^2$ value | Time Window |
| --- | --- | --- | --- | --- |
| North America | 0.0000000052 | 0.0082 | 0.98 | '2022-03' - '2022-05' |
| South America | 0.0028071909 | 0.0554 | 0.99 | '2021-12' |
| Europe | 0.0000376295 | 0.0099 | 0.95 | '2022-02' - '2022-03' |
| Oceania | 0.0000000000 | 0.0064 | 0.99 | '2022-01', '2022-02', '2022-03', '2022-04' |
| Asia | 0.0001752538 | 0.0577 | 0.99 | '2021-12' |
| Africa | 0.0000328541 | 0.0351 | 0.93 | '2021-11' - '2021-12' |

**Table S6:**
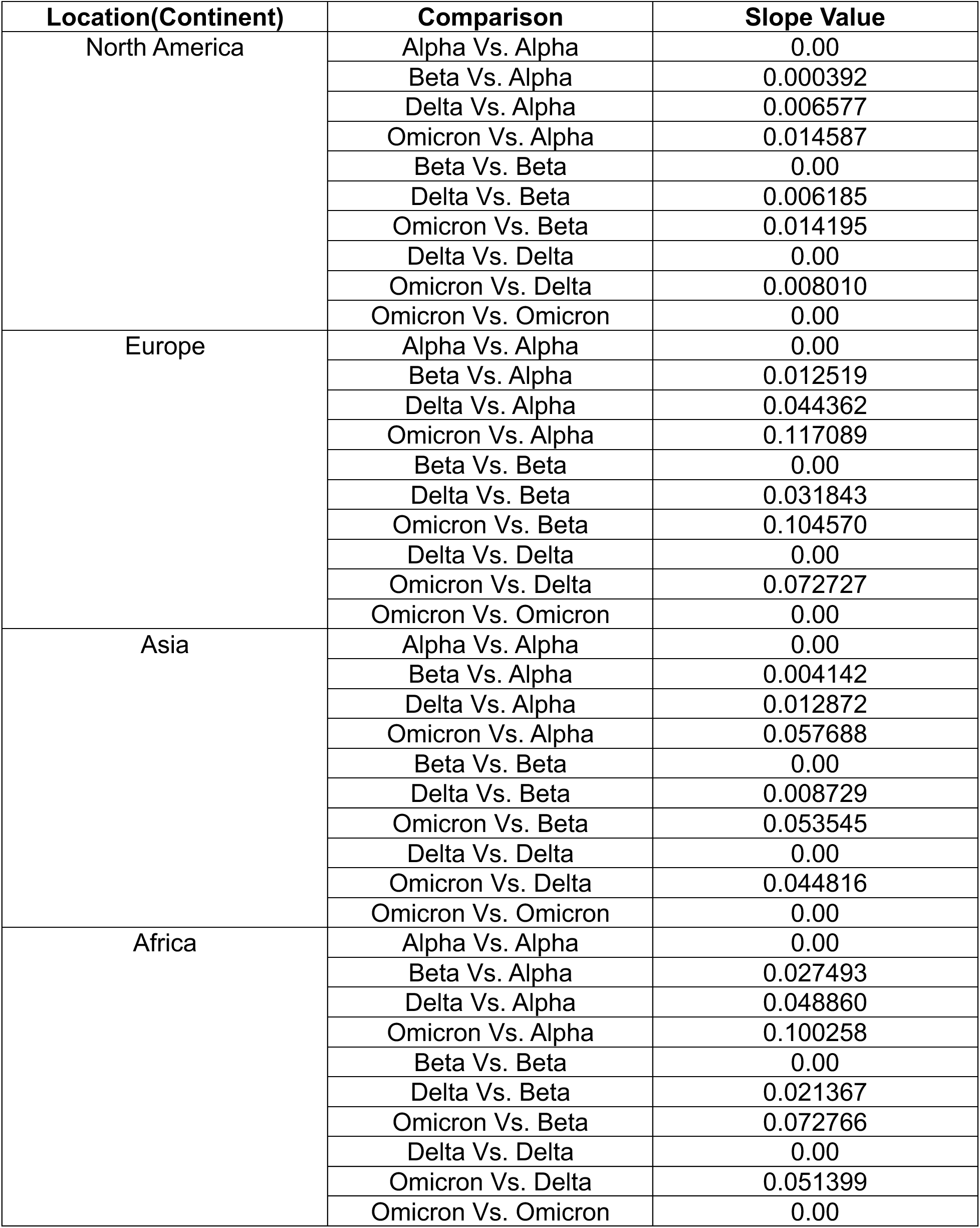
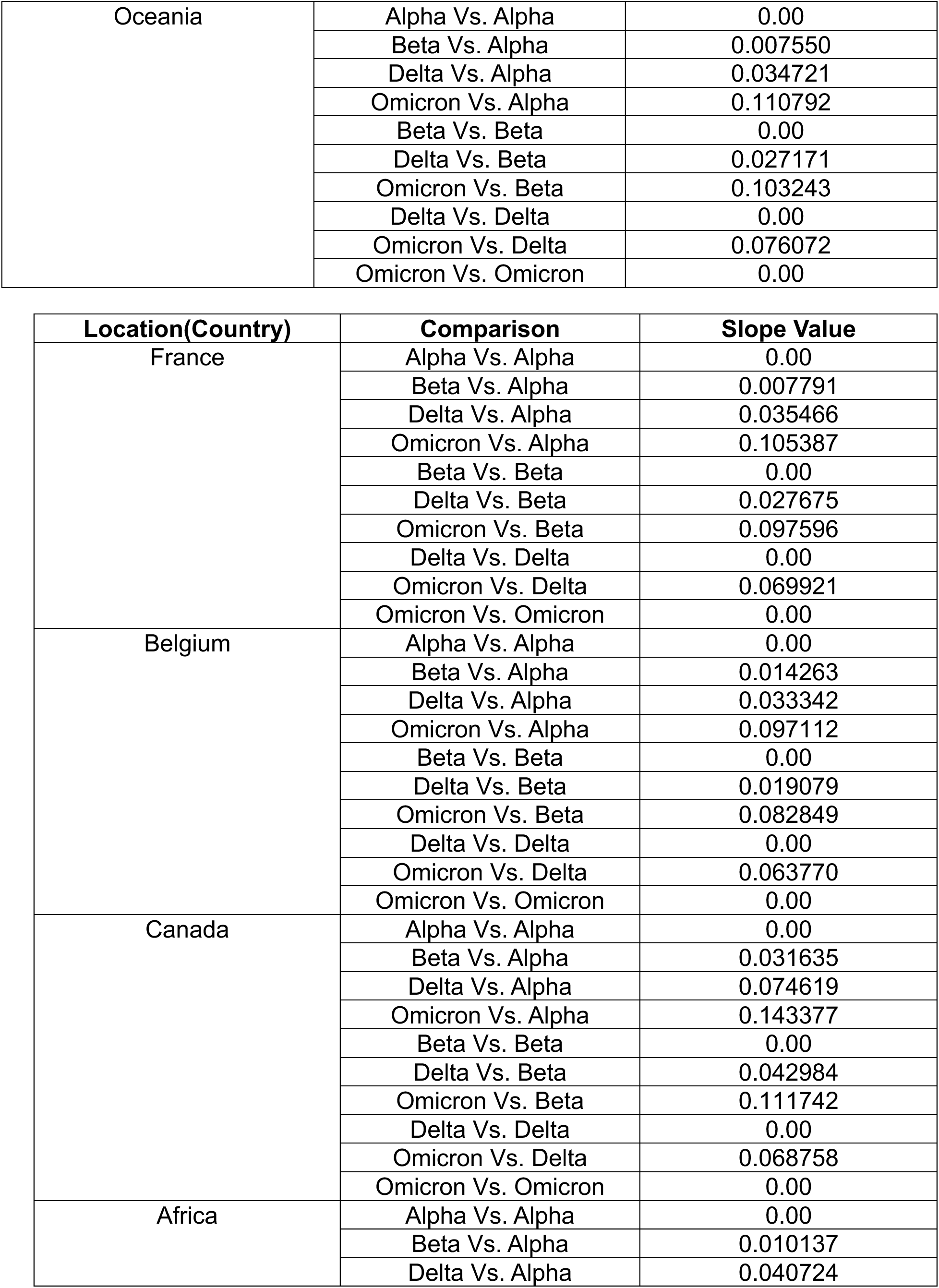
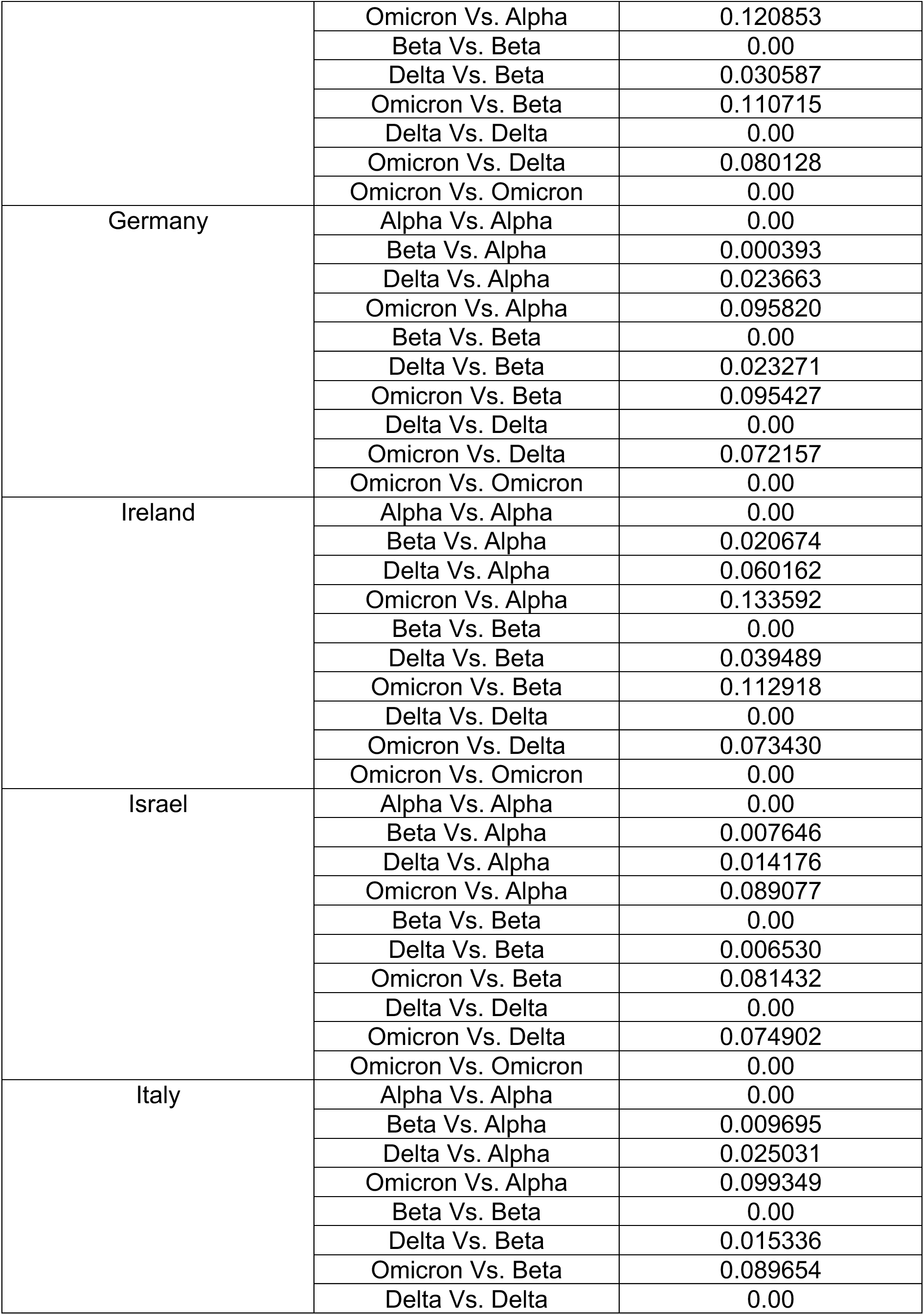
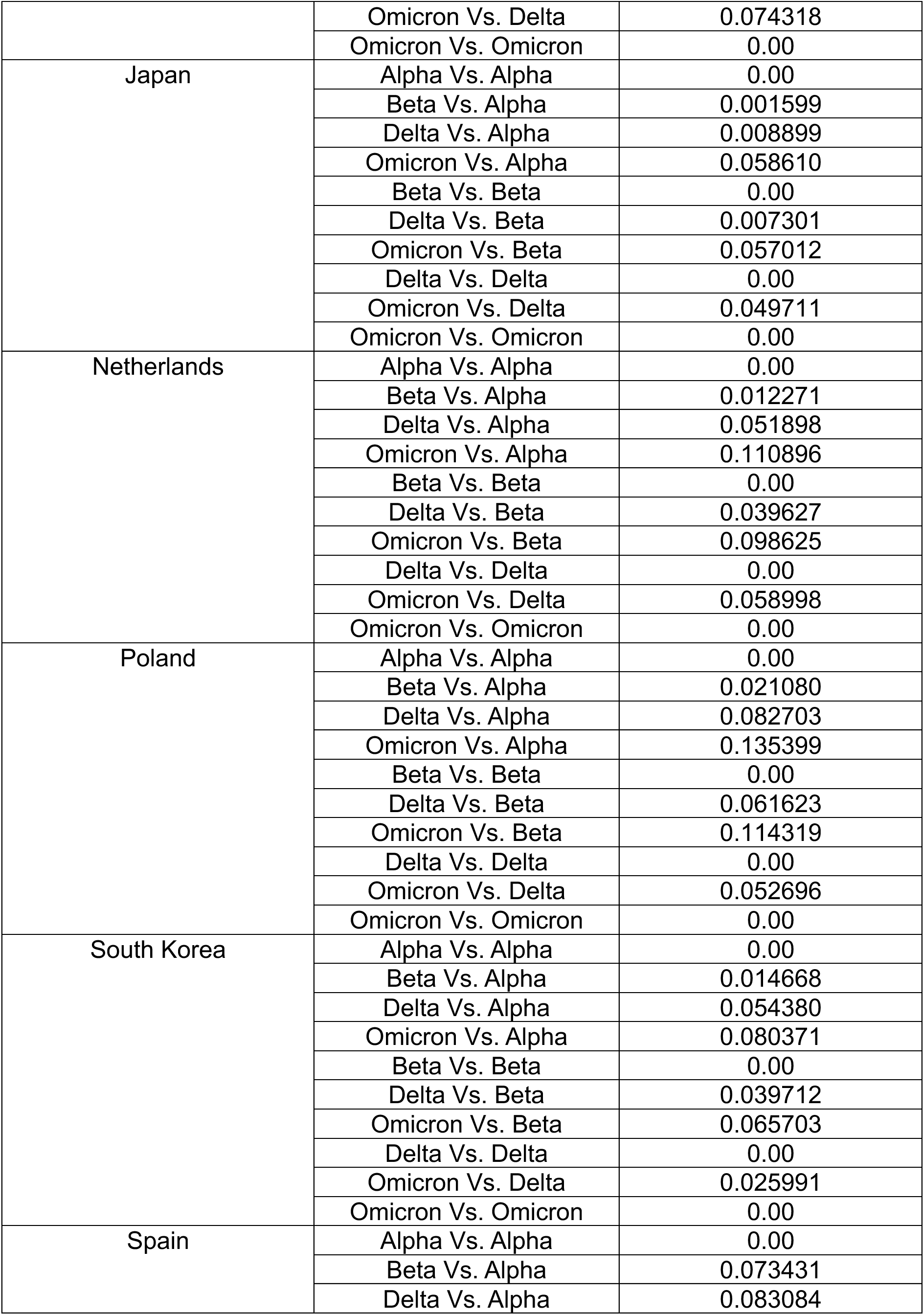
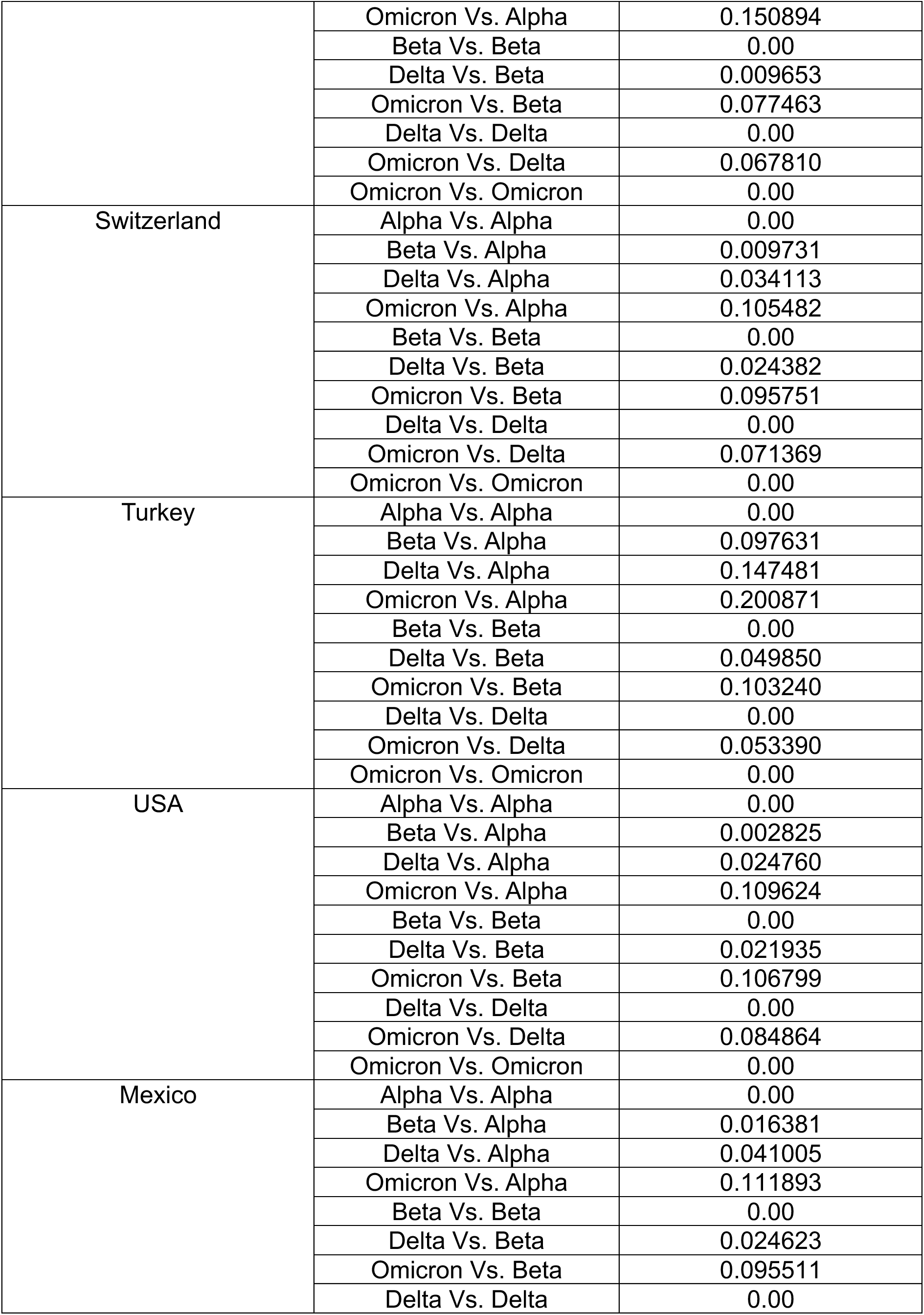
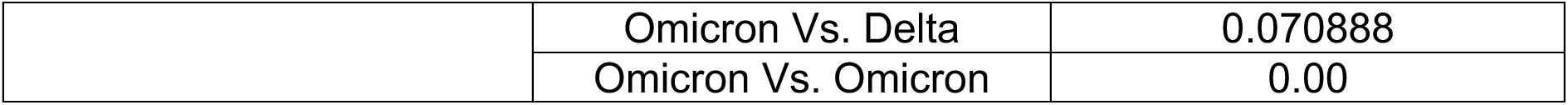
DPGR Estimates for All Variant Pairs (WHO Label)

## Notes

### Competing Interest Statement

The authors have declared no competing interest.

